# The MobiliseMe study: A randomised controlled efficacy trial of a cognitive behavioural therapy smartphone application (ClearlyMe®) for reducing depressive symptoms in adolescents

**DOI:** 10.1101/2024.11.17.24317363

**Authors:** Bridianne O’Dea, Sophie H. Li, Mirjana Subotic-Kerry, Melinda R. Achilles, Andrew J. Mackinnon, Philip J. Batterham, Helen Christensen, Anna Roberts, Komal Nagendraprasad, Zoe Dudley, Benjamin Gillham, Aliza Werner-Seidler

## Abstract

**Background:** The effectiveness of Digital Cognitive Behavioural Therapy (dCBT) smartphone applications for reducing depressive symptoms in adolescents remain unclear.

**Methods:** An online three-arm, parallel-group randomised controlled trial evaluated the effectiveness of a CBT smartphone application (ClearlyMe^®^) for reducing depressive symptoms in adolescents with outcomes assessed at baseline, post intervention (primary endpoint: 6-weeks post baseline) and follow-up (secondary endpoint: 4-months post baseline). The University of New South Wales Human Research Ethics Committee provided ethical approval. Youth were eligible if they were aged 12 to 17 years, in Australia, had mild to moderate depressive symptoms as measured by the adolescent Patient Health Questionnaire-9 (PHQ-A), were not receiving treatment or experiencing recent or severe suicidality, had access to a smartphone, and parental consent. Participants were randomised to self-directed ClearlyMe^®^, ClearlyMe^®^ with SMS-guided support, or the attention-matched control. Participants were not directly informed of their allocation. The statistician was blinded for analysis. The primary outcome was PHQ-A change post intervention. Intention-to-treat analyses used mixed models for repeated measures. The trial was prospectively registered on the Australian New Zealand Clinical Trials Registry (ACTRN12622000131752).

**Outcomes:** 569 adolescents (Mean age: 15.89, SD: 1.26, 74.2% female) were included in the analyses. The self-directed and guided conditions showed significantly greater reductions in depressive symptoms post intervention than the control (self-directed: Cohen’s *d*=0.35, mean differential decline 1.77; 95%CI: 0.56 – 2.98; *P*=.004; guided: *d*=0.33, mean differential decline: 1.31; 95%CI: 0.12 – 2.49; *P*=.030). The effects of self-directed and guided were comparable. Effects were also more robust and substantially larger post intervention among adolescents with probable MDD at baseline. Secondary outcomes showed similar patterns of change, although no differential effects for anxiety. There were no differences between the conditions at follow-up for any outcomes. Risk of adverse events was almost double in controls compared to self-directed (IRR: 1.73, 95%CI: 1.15 – 2.62, *P*=.009) and guided (IRR: 1.98 (95%CI: 1.27 – 3.08, *P*=.002).

**Interpretation:** ClearlyMe^®^, self-directed or with SMS-guided support, was effective for the short-term reduction of depressive symptoms in adolescents who have mild to moderate depression and are not receiving any other treatment.

*Funding:* The Goodman Foundation and the Australian National Health and Medical Research Council Investigator Grants (MRF1197249, GNT2008839, GNT115614).

## Introduction

Up to one third of adolescents (10 to 19 years) worldwide experience elevated depressive symptoms and nearly one in ten have Major Depressive Disorder (MDD) ^1^. Adolescent depression negatively impacts health, education, employment, and social functioning ^2–4^. Self-directed Digital Cognitive Behavioural Therapy (dCBT) is an evidence-based psychological treatment that can offer immediate and private support ^5,6^. Traditionally, most dCBT has been delivered through web-based computer programs, with small but significant effects on depressive symptoms ^7–10^. However, uptake of these programs has been limited, prompting a shift towards smartphone-based interventions.

Smartphone applications (apps) may improve the accessibility of dCBT, yet variations in design and user experience may influence engagement and treatment effectiveness ^11^. While mental health apps have demonstrated feasibility, few trials have confirmed the efficacy of CBT-based apps for reducing depressive symptoms in adolescents ^12–16^. A meta-analysis of 12 trials involving adolescents reported small but significant effects of apps for reducing internalising symptoms, with CBT being the most common therapeutic approach ^17^. However, another meta-analysis of 36 trials for adolescents found uncertain evidence for depression-specific apps, with only two CBT-based apps showing short-term benefits ^18^. Similarly, a review of 12 trials concluded the evidence supporting smartphone apps for adolescent depression remains insufficient ^19^. Many studies lack long-term follow-up and few of the tested apps have been publicly disseminated ^20^. These findings suggest that existing smartphone apps are suboptimal for adolescent depression ^19^.

Despite the widespread availability of mental health apps, engagement remains low. Over half of the top-searched mental health apps report no active users ^21^ and CBT-based apps have particularly low market reach compared to mindfulness apps ^22^. This may be due to limited adolescent involvement in co-design ^23,24^ alongside the motivational deficits associated with depression such as anhedonia, fatigue and impaired perceptions of value and reward ^25^.

Additionally, the role of guided support in enhancing engagement and treatment efficacy for adolescents remains underexplored. Most previous trials have evaluated self-directed apps ^20,26^, yet systematic reviews suggest that human-support can improve adherence and outcomes in digital mental health interventions ^27^. The effectiveness of guided support in smartphone-based dCBT for adolescent depression is therefore unclear.

This trial evaluated the effectiveness of a CBT-based smartphone app (ClearlyMe^®^) for reducing adolescent depressive symptoms, comparing self-directed and guided use with an attention-matched control. We hypothesised that participants receiving ClearlyMe^®^ (self-directed or guided) would experience greater reductions in depressive symptoms between baseline to 6-weeks (primary endpoint) and four months (secondary endpoint) compared to the control. We assessed clinical importance by examining changes in probable clinical caseness of MDD. This trial also examined the secondary effects of ClearlyMe^®^ on anxiety, psychological distress, emotional well-being, quality of life, rumination, emotion regulation, and CBT skill acquisition. We expected greater improvements in these outcomes among ClearlyMe^®^ users (both self-directed and guided). Engagement was also assessed, with the hypothesis that guided support would enhance app use. Additionally, we explored whether treatment effectiveness varied by baseline depression severity, gender, and openness to smartphone delivered therapies. Findings will inform the potential for ClearlyMe^®^ to deliver first-line psychological treatment to adolescents without clinician support.

## Methods

### Study design

A three-arm, parallel-group randomized controlled trial (RCT) with an equal allocation ratio. Reporting follows the CONSORT checklist for RCTs ^28^ and extension for E-Health trials ^29^. Outcomes were assessed at baseline, post intervention (primary endpoint: 6-weeks post baseline), and follow-up (secondary endpoint: 4-months post baseline). The trial sponsor was University of New South Wales (UNSW) Sydney. Ethical approval was obtained from UNSW Human Research Ethics Committee (HC#210889). The protocol followed SPIRIT guidelines and was published prior to recruitment ^30^. The trial was prospectively registered with the Australian New Zealand Clinical Trials Registry (ACTRN12622000131752) and allocated a UTN U1111-1271-8519. All participants reached the primary endpoint by May 2023, and the secondary endpoint by July 2023.

### Involvement of Lived Experience

Youth with Lived Experience (YLE) were involved in the design of the ClearlyMe app, creating content for inclusion in the app, alongside the study design, review of protocol, and co-design of recruitment materials. Many of the authors are also working from a position of Lived Experience.

### Participants

Participants were Australian-residing adolescents aged between 12 to 17 years old who had access to a smartphone and the Internet; an email address and mobile phone number; could read English at Grade 7-8 level; consent from a parent or guardian; and self-reported mild to moderately-severe depressive symptoms at screening (i.e., total score on the Patient Health Questionnaire-9 for Adolescents [PHQ-A] ranged from 5 to 19). Participants were excluded if they were receiving or planning professional psychological treatment for depression or anxiety; taking or beginning prescribed medication for depression or anxiety; reported severe suicidal ideation in past two weeks (scoring ≥2 on item 9 of the PHQ-A, which asks about frequency of thoughts of being better off dead or of hurting one’s self) or had serious thoughts or intentions of self-harm or suicide attempts within the past month at screening.

### Sample Size

The required sample size was 489 based on α=0.05, power=0.8, small to medium within-group effect sizes for the conditions (psychoeducation d=0.10)^31^, self-directed CBT (d=0.35) ^32^, and guided CBT (d=0.65) ^8,9^, and an attrition rate of 20% between baseline and primary endpoint.

### Randomisation and blinding

Randomisation followed the International Council for Harmonisation guidelines ^33^. Randomisation occurred after baseline using an automated computerised randomisation procedure within the Black Dog Institute Research Engine. A stratified randomization approach (1:1 ratio) with a block size of 6 ensured balance across age (12 to 14 years vs. 15 to 17 years) and symptom severity score on PHQ-A (≤ 9 vs. ≥10). Participants were not directly informed of their allocation (i.e., control versus active interventions) but may have deduced it from differences across conditions described in the participant information form. The statistician and chief investigators were blinded to allocations for the primary outcome analyses. Staff who provided the guided support or trial safety procedures were unblinded to allocations but were not involved in data analysis.

### Recruitment

Recruitment was active between May 2022 and March 2023 and consisted of social media advertisements on Facebook, Instagram, TikTok and Snapchat (83%), Black Dog Institute website (6%), word of mouth (13%) and unspecified (4%).

### Procedure

The trial was conducted online. Advertisements directed adolescents to the study webpage to complete the screener. Eligible participants provided consent via the Adolescent Participant Information and Consent Form (A-PICF), while ineligible participants received mental health service information. Upon consent, adolescents registered with the Black Dog Institute Research Engine, providing personal details. They then received the Parent Participant Information and Consent Form (P-PICF), requiring a parent’s contact details and electronic signature within a week. Three reminders were sent, and researchers followed up on any invalid emails. Consented adolescents were completed the baseline assessment within 7 days, those who failed to do so were withdrawn. After baseline, participants were randomised and received intervention instructions via email and Short Message Service (SMS). At primary and secondary endpoints, participants had days to complete study assessments (∼20 minutes) on any Internet device. They received two reminders and were reimbursed 10AUD for each assessment (up to 30AUD). Guided-condition participants received an additional 25AUD at randomisation for SMS costs. Participants were instructed to use the app’s Get Help section for mental health support, which was not monitored by staff. No further contact was provided unless technical support was requested.

### Conditions

Self-directed condition: ClearlyMe^®^ is a free, self-directed CBT-based smartphone app designed to provide therapeutic content and self-management strategies for depression.

Developed by researchers at the Black Dog Institute through a human-centred co-design process with adolescents, clinicians, and parents ^34^, the app delivers CBT content through 37 ultra-brief activities (<10 minutes each), grouped 9 psychological themes (i.e., collections). Each collection takes approximately 20 minutes to complete. Other features include mood tracking (MoodCheck), coping strategies (MindHacks), and lived experience video stories. Engagement tools included in-app reminders, saving and favouriting functions. A “Get Help” section linked users to external mental health support services and information (see Supplementary Material 1). Participants were instructed to complete one collection per week for 6 weeks in any order, receiving one weekly SMS reminder. In-app notifications prompted reengagement after 7 days of inactivity, but only for those who enabled them. Participants who failed to download the app within 7 days of randomisation received one email and one SMS reminder.

Guided support condition: Participants in this condition were provided with the same instructions and expectations for use and app access as the self-directed condition. However, these participants also received a weekly guided support chat session via SMS. All chats were delivered by non-clinical research staff using a standard script and decision flow chart. The chats focused on adherence (i.e. completion of one collection per week) by providing technical support and low-intensity motivational coaching to overcome barriers to use. Chats occurred between 3:30pm and 7pm Mondays to Thursday AEST and were overseen by clinical psychologists. Staff made one additional contact attempt within two days if there was no response and all weekly chats were initiated regardless of prior responsiveness. All chats were stored and reviewed for script adherence.

Control condition: The active attention-matched control condition consisted of weekly digital psychoeducation Portable Document Formats (PDFs) delivered by SMS. Content was adapted from interventions used in past adolescent mental health trials ^31^ and focused on common mental health problems, self-care activities, and links to Australian mental health organisations. Participants in this condition were granted access to ClearlyMe^®^ after completion of the 4-month follow-up.

### Primary Outcome

Depressive symptoms were measured by total scores (range: 0 to 27) on the 9-item self-report PHQ-A, which has been validated for use in adolescents. This no-cost brief measure takes less than 5 minutes to complete and has been widely used across clinical, community, and at-risk settings for youth. Probable cases of Major Depressive Disorder (MDD) are indicated by the total score cut point of ≥15, which aligns with moderately severe to severe symptoms and a need for treatment.

### Secondary Outcomes

#### Generalised anxiety symptoms

Assessed using total scores (range: 0 to 21) on the 7-item self-report Generalized Anxiety Disorder-7 scale (GAD-7), which has been validated for use in adolescents (trial α = .79).

#### Psychological Distress

Assessed using total scores (range: 5 to 25) on 5-item self-report Distress Questionnaire-5 (DQ-5), which has been validated to assess the frequency of psychological distress in adolescents in the past 30 days (trial α = .70) ^35^.

#### Emotional wellbeing

Assessed using total scores (range: 7 to 35) on the 7-item self-report Short Warwick-Edinburgh Mental Wellbeing Scale (SWEMWBS), which has been validated to assess adolescents’ wellbeing in the past 14 days (trial α = .73).

#### Quality of life

Assessed using total scores (range: 9 to 45) on the 9-item self-reported Child Health Utility 9D (CHU-9D), which has been validated to assess health-related quality of life in adolescents across nine domains: worry, sadness, pain, tiredness, annoyance, schoolwork/homework, sleep, daily routine, and ability to join activities (trial α = .77).

#### Rumination

Assessed using the 10-item self-report Ruminative Responses Scale – short version (RRS), which is validated to assess how adolescents typically react when feeling down, sad or depressed. The scale comprises two subscales: reflection and brooding. Items were summed to generate two sub-totals and an overall score (range: 10 to 40, trial α =.74).

#### Emotion regulation

The 10-item Emotion Regulation Questionnaire for Children and Adolescents (ERQ-CA) evaluated the use of cognitive reappraisal (6-items) and expressive suppression (4-items) as emotion regulation strategies. Subscale items were summed to produce a cognitive reappraisal (ERQ-CA-CR) score (range: 6 to 30, trial α = .83) and expressive suppression score (ERQ-CA-ES) (range: 4 to 20, trial α = .67). Higher scores indicate greater utilisation of each strategy.

#### CBT skill acquisition

Assessed using the 16-item Cognitive Behavioural Therapy Skills Questionnaire (CBTSQ), which consisted of two subscales: cognitive restructuring (CBTSQ-CR; 9-items, range: 9 to 45) and behavioural activation (CBTSQ-BA; 7-items, range: 7 to 35). Higher scores indicate greater utilisation of CBT skills (trial α = .84).

#### Engagement with ClearlyMe^®^

Uptake was measured by the proportion of participants who downloaded the ClearlyMe^®^ app. Adherence was measured by the number of collections (out of 9) completed within six weeks. The number of lessons completed (out of 37) was also measured. The 6-item self-reported Digital Working Alliance Inventory (DWAI) assessed three core domains of digital technology use (goals, tasks and bond) across the two intervention conditions at post intervention only. Items were summed to generate a total score (range: 6 to 42) with higher scores indicating greater alliance (trial α =.88).

#### Program satisfaction and barriers to use

Assessed post intervention using a purpose-built 11-item measure of participants’ satisfaction, experience and perceived helpfulness of their allocated intervention ^31,36^. The first 8 items prompted participants to agree or disagree with statements on satisfaction and experience with the intervention. Participants also rated the overall helpfulness of their intervention on a 5-point Likert scale. Two free response questions examined the helpful aspects of the digital program (e.g., In what ways did the intervention help you?) and suggestions for improvement (e.g., What would make the intervention better?). The Digital Program Barriers questionnaire ^31,36^ assessed participants’ experiences of various barriers to the allocated interventions post intervention, including technical issues, accessibility, and individual factors (answered ‘yes’ or ‘no’).

#### Background characteristics

Age, gender (female, male, non-binary, different identity, free response option), Aboriginal or Torres Strait Islander identity (yes, no, prefer not to say) and/or Lesbian, Gay, Bisexual, Transgender, Queer and/or Intersex identity (yes, no, I’d prefer not to say), home state or territory, categorical descriptor of home location (metropolitan, regional, or rural/remote), and school grade. Participants were asked whether they had (i) been diagnosed with depression and/or anxiety by a health professional, (ii) received psychological treatment for a mental health problem or mental illness from a health professional, (iii) taken prescribed medication for a mental health problem or mental illness like depression or anxiety, (iv) felt like they needed help for a mental health issue like depression or anxiety. Participants were asked to report whether they have ever used any mobile app to help with their emotional wellbeing or mental health (yes, no, I’d rather not say), and if yes, whether they found it helpful (yes, no). Participants were asked to rate the extent to which they felt their emotional wellbeing could be improved by a mobile app (referred to as ‘openness to smartphone applications for emotional wellbeing’) using a 5-point scale ranging from not at all (1) to extremely (5). This variable was then dichotomised for analysis: ‘moderate to high’ (scores > 2) vs. ‘low’ (scores < 3).

### Data collection and analysis

Trial data was collected on the Black Dog Institute Research Engine. Analyses were conducted using intention-to-treat approach and included all participants regardless of intervention uptake or use. Missing due to drop out or any other factor was assumed to be missing at random. Linear Mixed Model for Repeated Measures (MMRM), with fixed factors of group and time and their interaction were used. The stratification factor of age group was tested in preliminary modelling but subsequently omitted for simplicity of interpretation as it had no effect on the results. Within person associations were accommodated using an unstructured variance-covariance matrix. The statistical significance of the ClearlyMe^®^ conditions were evaluated in a planned comparison of change from baseline to post intervention compared to change in the control group. The Kenward-Roger method, using observed information matrix, was used to adjust degrees of freedom for tests. Effect sizes were estimated post intervention and at follow-up using differences in estimated marginal means and the model residual variance. Mixed logistic regression with a random participant intercept were used to model changes in probable MDD cases post intervention and follow-up. Comparable methods were used to evaluate differential change at follow-up and for outcomes of secondary variables. The Benjamini-Hochberg method was used for post hoc adjustments to the P value. Separate exploratory analyses examined treatment effects in participants with probable MDD at baseline, males vs. females, and those with low vs. high openness to apps for emotional wellbeing. Engagement was assessed using models exploring condition effects on collections and activities completed. A zero-inflated negative binomial model with a constant inflation component best fit activity completion data. A series of models of increasingly complexity explored the relationship between ClearlyMe^®^ condition (self-directed vs guided) on the number of collections completed due to substantial percentage of participants completing nil. A zero-inflated negative binomial model with both the count and zero inflation components predicted by group condition fitted the data better than Poisson and negative binomial models without zero inflation or with constant inflation The harms analysis used descriptive statistics and logistic regressions with relative risk ratios. Analysis was undertaken using Stata v18.

## Results

Shown in Figure 1, 665 adolescents completed baseline and were allocated. A total of 569 were retained in the final baseline sample. Reasons for removal included fraudulent or duplicate responses, technical errors, or active withdrawals (see Figure 1). A total of 75.6% of the baseline sample (*n=*430/569) were retained at post intervention and 62.0% retained at follow up.

**Figure 1.**
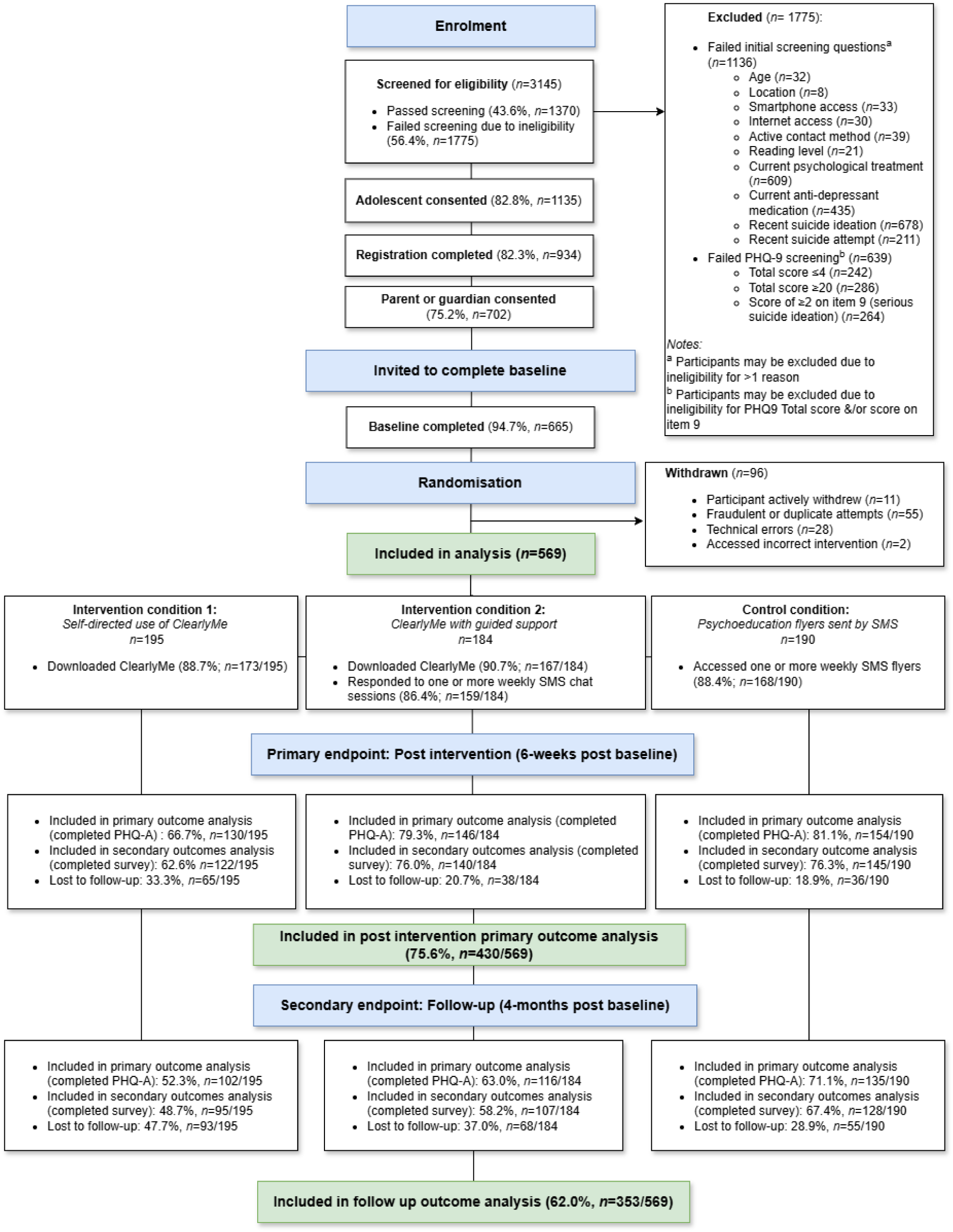
CONSORT diagram showing the flow of participants in the MobiliseMe trial.

Table 1 outlines the participants’ characteristics at baseline. In the final sample, the majority were female (*n=*422/569; 74.2%), aged 15-17 years (*n=*477/569; 83.8%) and currently attending secondary school (*n=*536/569; 94.2%). For missingness, being older, or living in a rural or remote location was associated with missing assessments (Relative Risk (RR) per year of age =1.15, *P* =.032, 95% CI:1.01 - 1.30, and RR=1.56, *P* =.039, 95% CI:1.02 - 2.39 respectively). Higher baseline scores on the ERQA-CA and CBTSQ-CR subscales reduced risk of missing assessments (RR per unit=0.96, *P*=.015, 95% CI:0.93 - 0.99 and RR per unit =0.97, *P* =.004, 95% CI:0.95 - 0.99 respectively).

**Table 1.**
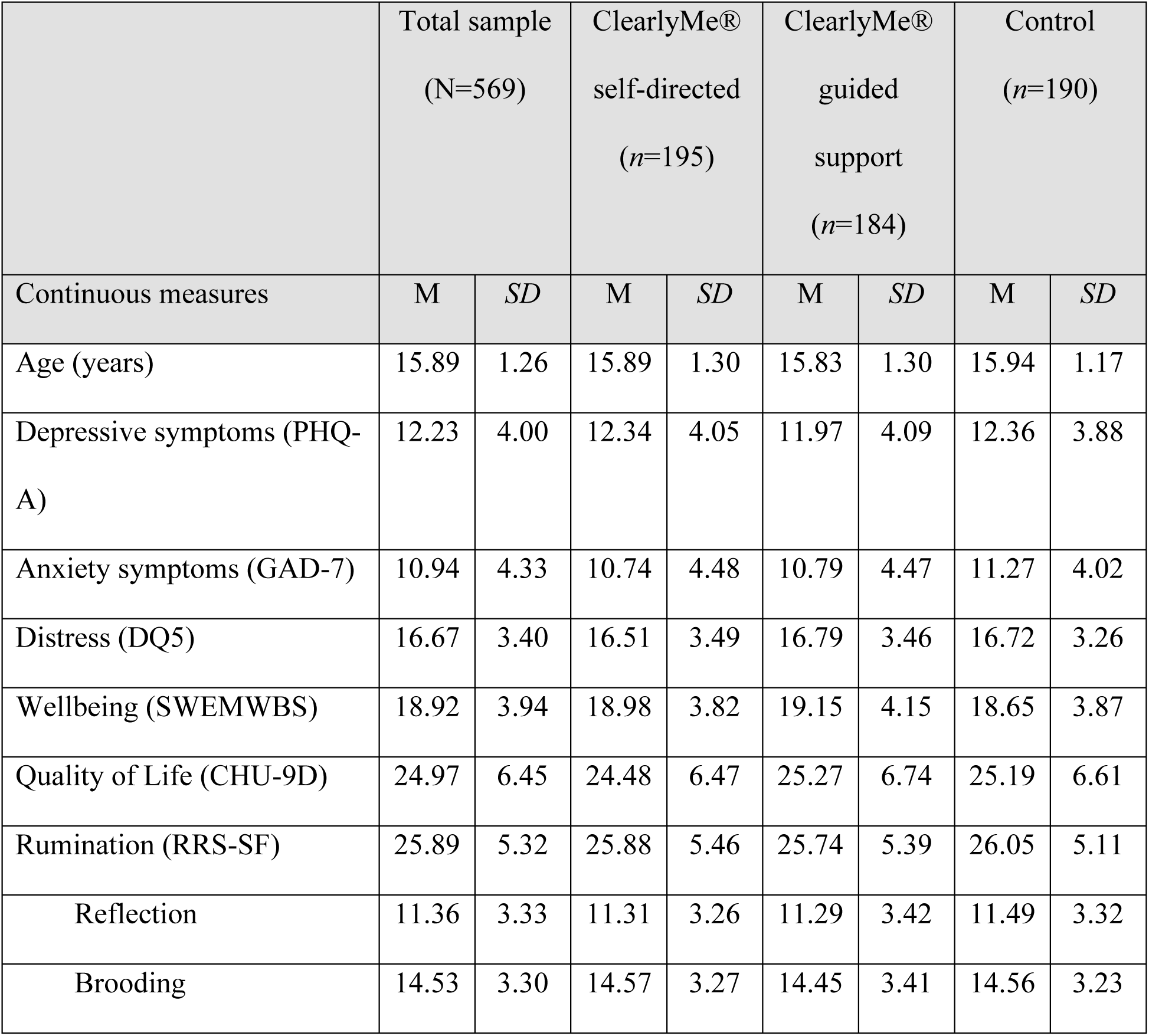
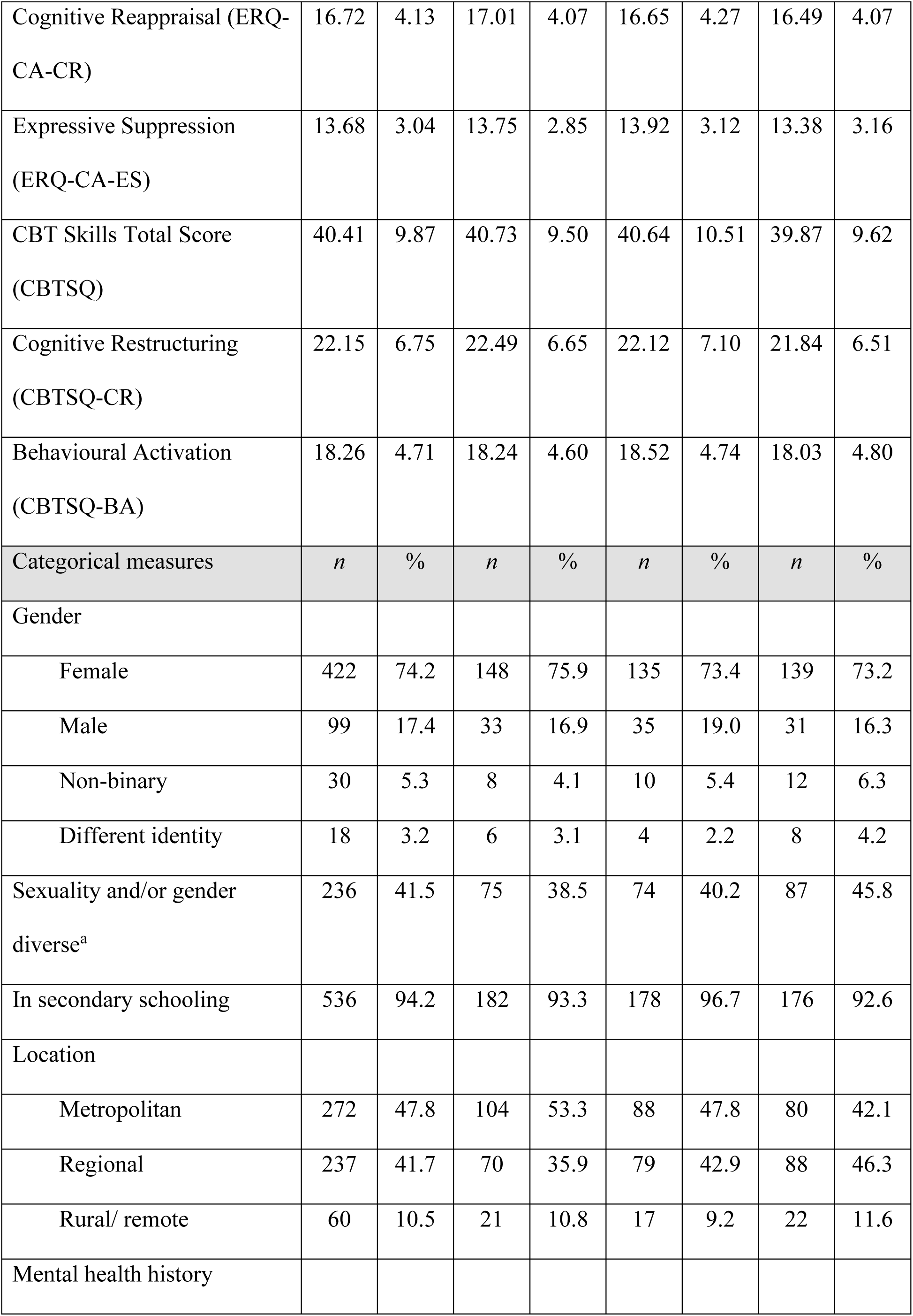
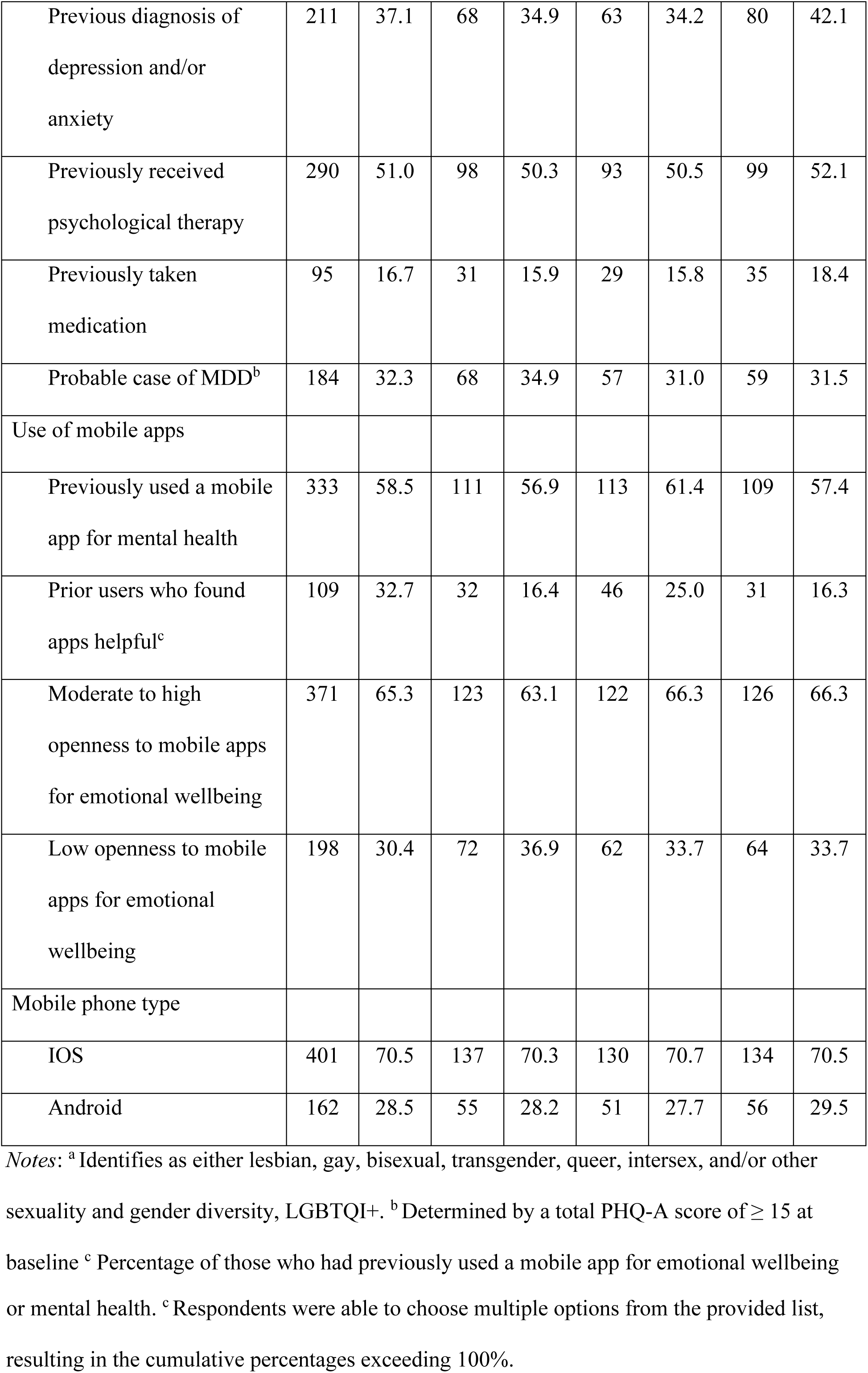
Participant characteristics at baseline (N=569).

|  | Total sample<br>(N=569) |  | ClearlyMe®<br>self-directed<br>(n=195) |  | ClearlyMe®<br>guided<br>support<br>(n=184) |  | Control<br>(n=190) |  |
| --- | --- | --- | --- | --- | --- | --- | --- | --- |
| Continuous measures | M | SD | M | SD | M | SD | M | SD |
| Age (years) | 15.89 | 1.26 | 15.89 | 1.30 | 15.83 | 1.30 | 15.94 | 1.17 |
| Depressive symptoms (PHQ-A) | 12.23 | 4.00 | 12.34 | 4.05 | 11.97 | 4.09 | 12.36 | 3.88 |
| Anxiety symptoms (GAD-7) | 10.94 | 4.33 | 10.74 | 4.48 | 10.79 | 4.47 | 11.27 | 4.02 |
| Distress (DQ5) | 16.67 | 3.40 | 16.51 | 3.49 | 16.79 | 3.46 | 16.72 | 3.26 |
| Wellbeing (SWEMWBS) | 18.92 | 3.94 | 18.98 | 3.82 | 19.15 | 4.15 | 18.65 | 3.87 |
| Quality of Life (CHU-9D) | 24.97 | 6.45 | 24.48 | 6.47 | 25.27 | 6.74 | 25.19 | 6.61 |
| Rumination (RRS-SF) | 25.89 | 5.32 | 25.88 | 5.46 | 25.74 | 5.39 | 26.05 | 5.11 |
| Reflection | 11.36 | 3.33 | 11.31 | 3.26 | 11.29 | 3.42 | 11.49 | 3.32 |
| Brooding | 14.53 | 3.30 | 14.57 | 3.27 | 14.45 | 3.41 | 14.56 | 3.23 |
| Cognitive Reappraisal (ERQ-CA-CR) | 16.72 | 4.13 | 17.01 | 4.07 | 16.65 | 4.27 | 16.49 | 4.07 |
| Expressive Suppression (ERQ-CA-ES) | 13.68 | 3.04 | 13.75 | 2.85 | 13.92 | 3.12 | 13.38 | 3.16 |
| CBT Skills Total Score (CBTSQ) | 40.41 | 9.87 | 40.73 | 9.50 | 40.64 | 10.51 | 39.87 | 9.62 |
| Cognitive Restructuring (CBTSQ-CR) | 22.15 | 6.75 | 22.49 | 6.65 | 22.12 | 7.10 | 21.84 | 6.51 |
| Behavioural Activation (CBTSQ-BA) | 18.26 | 4.71 | 18.24 | 4.60 | 18.52 | 4.74 | 18.03 | 4.80 |
| Categorical measures | *n* | % | *n* | % | *n* | % | *n* | % |
| Gender |  |  |  |  |  |  |  |  |
| Female | 422 | 74.2 | 148 | 75.9 | 135 | 73.4 | 139 | 73.2 |
| Male | 99 | 17.4 | 33 | 16.9 | 35 | 19.0 | 31 | 16.3 |
| Non-binary | 30 | 5.3 | 8 | 4.1 | 10 | 5.4 | 12 | 6.3 |
| Different identity | 18 | 3.2 | 6 | 3.1 | 4 | 2.2 | 8 | 4.2 |
| Sexuality and/or gender diverse<sup>a</sup> | 236 | 41.5 | 75 | 38.5 | 74 | 40.2 | 87 | 45.8 |
| In secondary schooling | 536 | 94.2 | 182 | 93.3 | 178 | 96.7 | 176 | 92.6 |
| Location |  |  |  |  |  |  |  |  |
| Metropolitan | 272 | 47.8 | 104 | 53.3 | 88 | 47.8 | 80 | 42.1 |
| Regional | 237 | 41.7 | 70 | 35.9 | 79 | 42.9 | 88 | 46.3 |
| Rural/ remote | 60 | 10.5 | 21 | 10.8 | 17 | 9.2 | 22 | 11.6 |
| Mental health history |  |  |  |  |  |  |  |  |
| Previous diagnosis of depression and/or anxiety | 211 | 37.1 | 68 | 34.9 | 63 | 34.2 | 80 | 42.1 |
| Previously received psychological therapy | 290 | 51.0 | 98 | 50.3 | 93 | 50.5 | 99 | 52.1 |
| Previously taken medication | 95 | 16.7 | 31 | 15.9 | 29 | 15.8 | 35 | 18.4 |
| Probable case of MDD<sup>b</sup> | 184 | 32.3 | 68 | 34.9 | 57 | 31.0 | 59 | 31.5 |
| Use of mobile apps |  |  |  |  |  |  |  |  |
| Previously used a mobile app for mental health | 333 | 58.5 | 111 | 56.9 | 113 | 61.4 | 109 | 57.4 |
| Prior users who found apps helpful<sup>c</sup> | 109 | 32.7 | 32 | 16.4 | 46 | 25.0 | 31 | 16.3 |
| Moderate to high openness to mobile apps for emotional wellbeing | 371 | 65.3 | 123 | 63.1 | 122 | 66.3 | 126 | 66.3 |
| Low openness to mobile apps for emotional wellbeing | 198 | 30.4 | 72 | 36.9 | 62 | 33.7 | 64 | 33.7 |
| Mobile phone type |  |  |  |  |  |  |  |  |
| IOS | 401 | 70.5 | 137 | 70.3 | 130 | 70.7 | 134 | 70.5 |
| Android | 162 | 28.5 | 55 | 28.2 | 51 | 27.7 | 56 | 29.5 |

Notes: <sup>a</sup> Identifies as either lesbian, gay, bisexual, transgender, queer, intersex, and/or other
sexuality and gender diversity, LGBTQI+. <sup>b</sup> Determined by a total PHQ-A score of $\geq 15$ at
baseline <sup>c</sup> Percentage of those who had previously used a mobile app for emotional wellbeing
or mental health. <sup>c</sup> Respondents were able to choose multiple options from the provided list, resulting in the cumulative percentages exceeding 100%.

Shown in Table 2 and Figure 2, depressive symptoms in the self-directed and guided conditions declined from baseline means by a significantly greater amount than the control condition post intervention, with small effects (*d*=0.33 to 0.35). At primary endpoint, both forms of ClearlyMe^®^ showed greater declines on the PHQ-A than did the control condition (self-directed vs control: mean differential decline 1.77; 95%CI: 0.56 – 2.98; *P*=.004. guided versus control: mean differential decline: 1.31; 95%CI: 0.12 – 2.49; *P*=.030). These changes remained significant after post hoc adjustment. Differential declines from baseline were not significant at follow-up. When examining the effects in the probable cases of MDD at baseline, the pattern of outcomes was generally the same as the whole sample. However, the significant effects were more robust and effect sizes substantially larger post intervention (*d*=0.65 for guided use vs control, *d*=0.71 for self-directed vs control).

**Figure 2.**
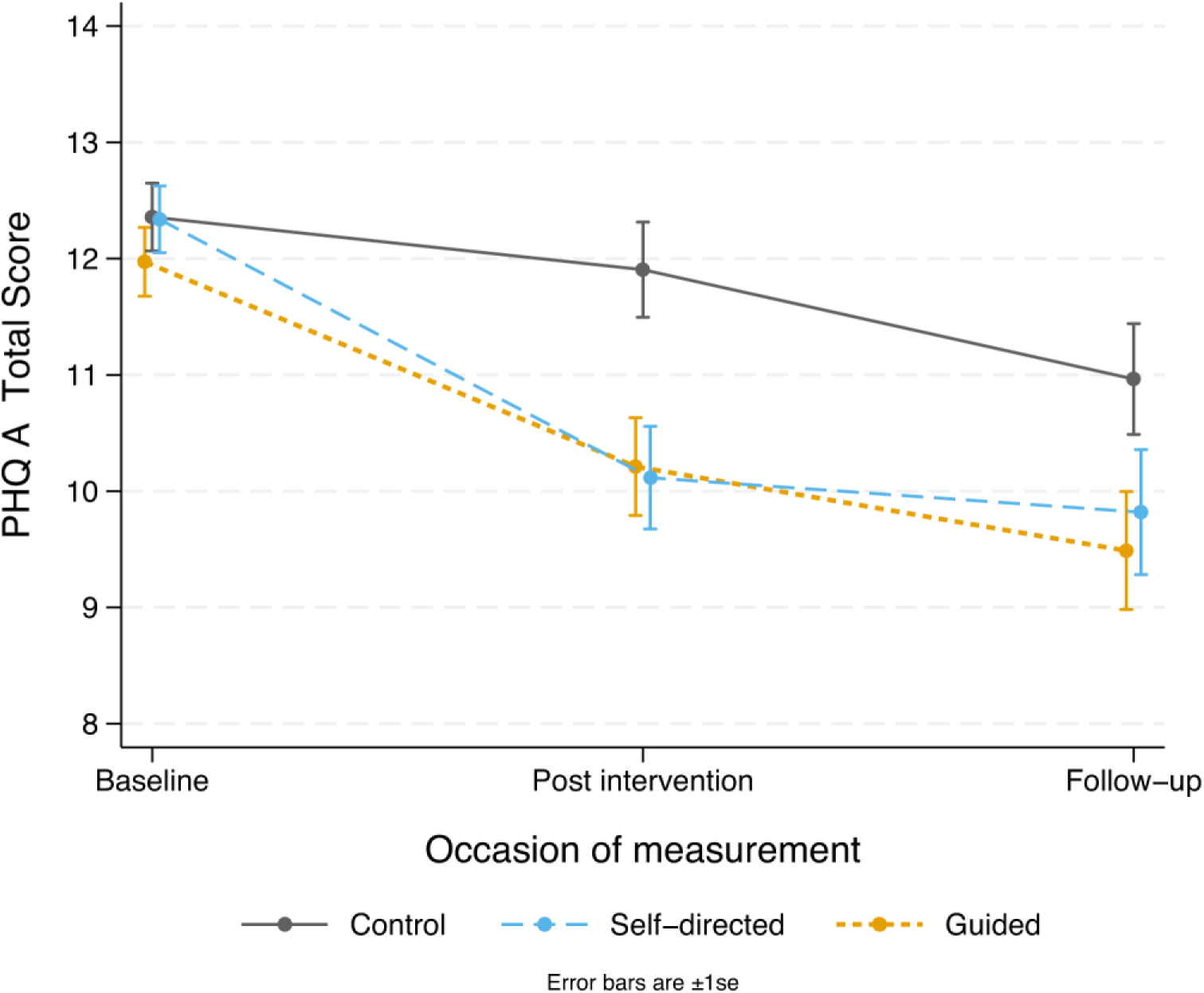
Estimated marginal means for depressive symptoms (PHQ-A, primary outcome) for the conditions on each occasion of measurement.

**Table 2.**
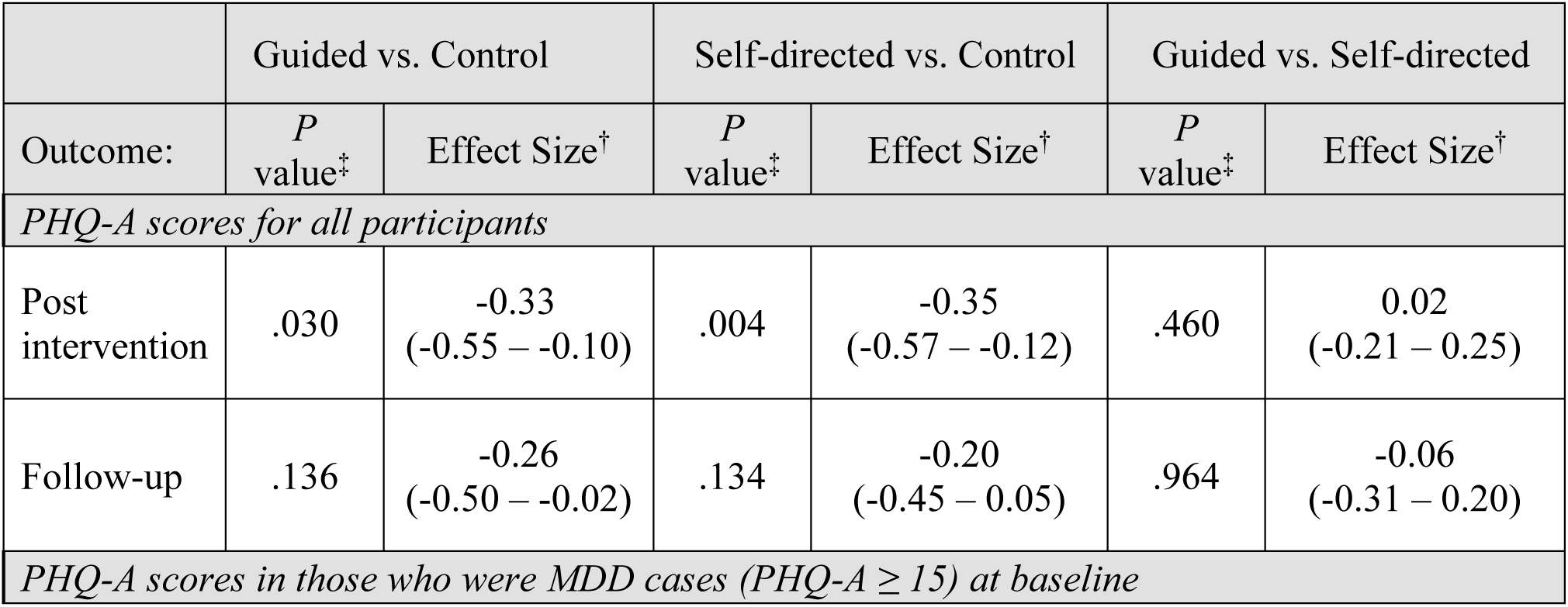
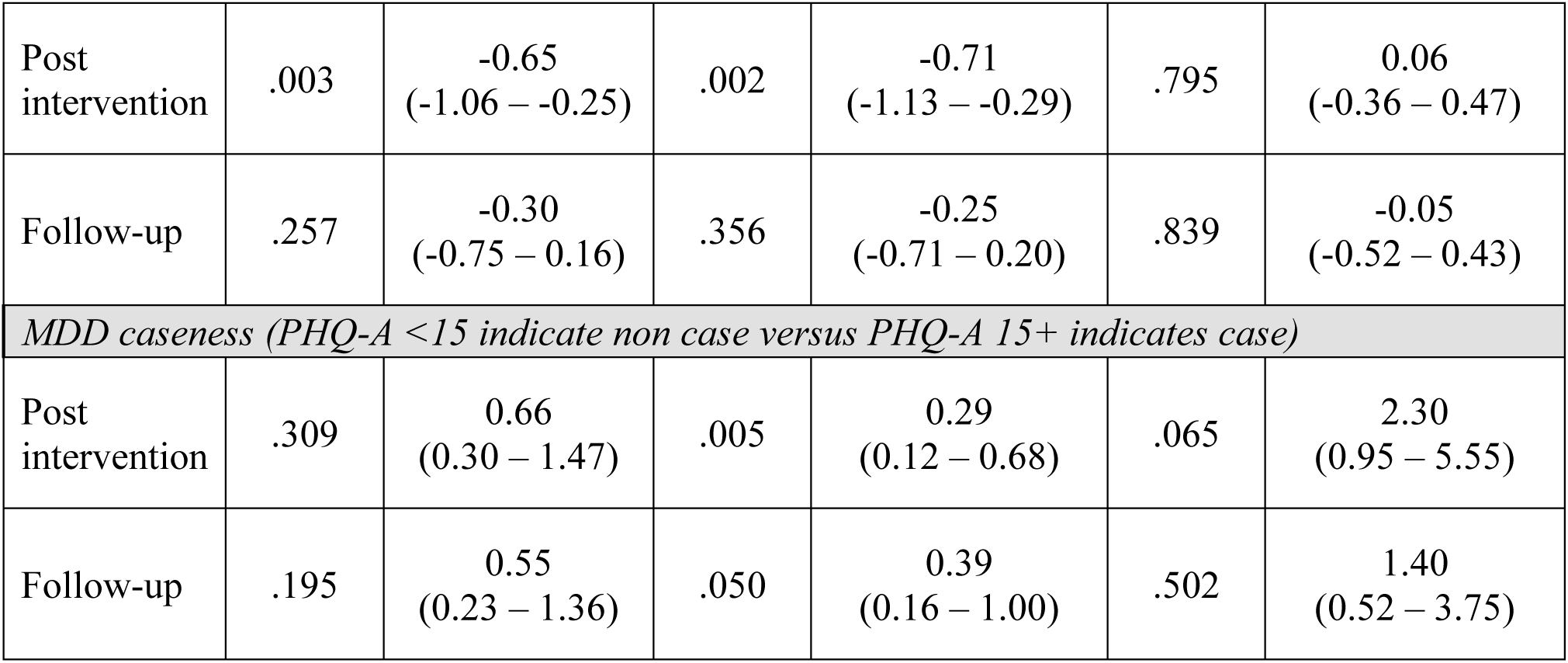
Tests of change in depressive symptoms (PHQ-A) and MDD cases from baseline to post intervention and follow-up and effect sizes.

|  | Guided vs. Control |  | Self-directed vs. Control |  | Guided vs. Self-directed |  |
| --- | --- | --- | --- | --- | --- | --- |
| Outcome: | $P$ value<sup>‡</sup> | Effect Size<sup>†</sup> | $P$ value<sup>‡</sup> | Effect Size<sup>†</sup> | $P$ value<sup>‡</sup> | Effect Size<sup>†</sup> |
| *PHQ-A scores for all participants* |  |  |  |  |  |  |
| Post intervention | .030 | -0.33<br>(-0.55 – -0.10) | .004 | -0.35<br>(-0.57 – -0.12) | .460 | 0.02<br>(-0.21 – 0.25) |
| Follow-up | .136 | -0.26<br>(-0.50 – -0.02) | .134 | -0.20<br>(-0.45 – 0.05) | .964 | -0.06<br>(-0.31 – 0.20) |
| *PHQ-A scores in those who were MDD cases (PHQ-A $\geq 15$) at baseline* |  |  |  |  |  |  |
| Post intervention | .003 | -0.65<br>(-1.06 – -0.25) | .002 | -0.71<br>(-1.13 – -0.29) | .795 | 0.06<br>(-0.36 – 0.47) |
| Follow-up | .257 | -0.30<br>(-0.75 – 0.16) | .356 | -0.25<br>(-0.71 – 0.20) | .839 | -0.05<br>(-0.52 – 0.43) |
| *MDD caseness (PHQ-A <15 indicate non case versus PHQ-A 15+ indicates case)* |  |  |  |  |  |  |
| Post intervention | .309 | 0.66<br>(0.30 – 1.47) | .005 | 0.29<br>(0.12 – 0.68) | .065 | 2.30<br>(0.95 – 5.55) |
| Follow-up | .195 | 0.55<br>(0.23 – 1.36) | .050 | 0.39<br>(0.16 – 1.00) | .502 | 1.40<br>(0.52 – 3.75) |

Due to elevated rates of attrition, a post hoc sensitivity analysis was undertaken, essentially assuming that participants with no observations at primary endpoint experienced being in the control group. Differential decline at endpoint was reduced in both interventions but remained significant for the self-directed version (*P*=.042) while the guided condition approached significance (*P*=.069). See Supplementary Material 2 (Figure S2) for more information.

The proportion of probable cases of MDD at primary endpoint was relatively stable in the control condition post intervention and the decline from baseline was not statistically significant (see Figure 3). The reduction in cases post intervention for the self-directed condition was substantially larger than for the guided condition. When compared to the control condition, the relative odds of change in cases post intervention were statistically significant for the self-directed condition but not the guided condition (see Table 2). This was, in part, attributable to the slightly higher baseline case rate in the self-directed condition. Proportions converged by follow-up and were not significantly lower than the control condition.

**Figure 3.**
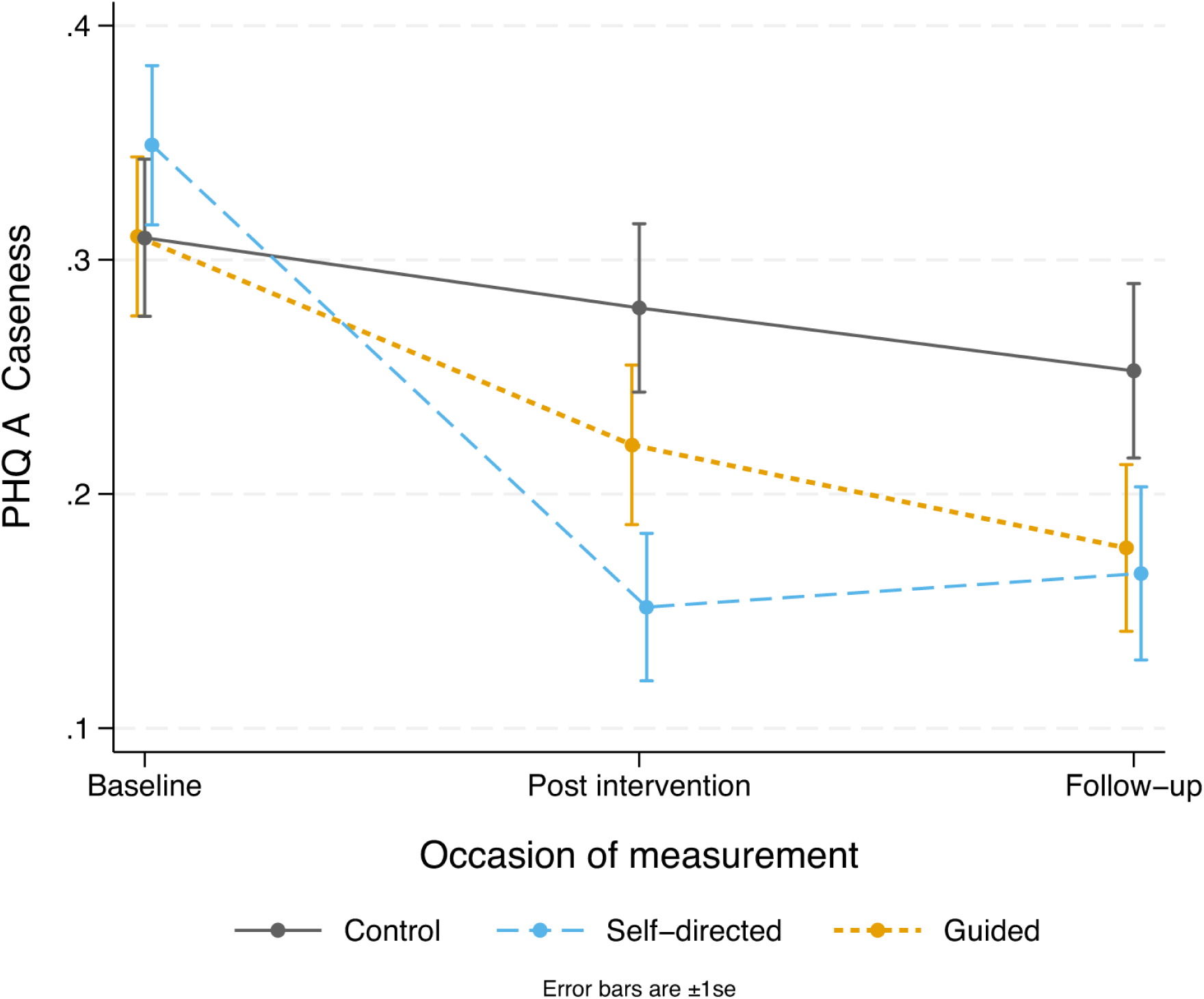
Predicted proportions of probable cases of Major Depressive Disorder (PHQ-A score ≥15) for the conditions on each occasion of measurement.

Expected adverse events (EAE; total of 171 events) were reported by 102 participants (17.9%). The relative risk of any EAE post baseline was almost double in the control condition compared to self-directed and guided with IRR of 1.73 (95%CI: 1.15 – 2.62, *P*=.009) and 1.98 (95%CI: 1.27 – 3.08, *P*=.002), respectively. Most of the EAEs (93, 54% of total events) were suicidal ideation with 77 participants reporting scores ≥ 2 on item 9 of the PHQ-A post baseline (13.5%). The relative risk of suicidal ideation in the control condition (20.5%) was more than double that of the self-directed condition (8.7%) and nearly double that of the guided condition (11.4%) with IRRs of 2.35 (95%CI: 1.38 – 4.02, *P*=.002) and 1.80 (95%CI: 1.10 – 2.94, *P*=.019), respectively. Mental health hospitalisations accounted for 18 EAEs (17.6%). There were 13 requests for a clinician call with 6 taken up. Only one participant who received ClearlyMe^®^ perceived that the app or trial activities had made their mental health worse.

The exploratory analysis for gender effects (males and females) on depressive symptoms yielded similar patterns as the whole sample (see Supplementary Material 2). However, there was low power for detecting gender effects given so few males in the sample (*n=*99). In participants with moderate to high openness to smartphone therapy at baseline, change in depressive symptoms at post intervention was significant in the self-directed and guided conditions (*P*=.009, *d*=0.34, 95%CI: 0.06 – 0.63 and *P*=.013. *d*=0.39, 95%CI: 0.11 – 0.66 respectively) but remained significant at follow-up in the guided condition only (*P*= .015, *d=*0.42, 95%CI: 0.12 – 0.71). In those with low openness at baseline, there were no significant changes in depressive symptoms at any time point in the self-directed or guided conditions when compared to the control (all *P* >.05), although, symptoms in the self-directed condition declined at follow-up in comparison to means in the guided condition, which converged to baseline (*P=*.051). See Supplementary Material 2 for Figures.

Shown in Table 3, changes in the secondary outcomes post intervention were comparable to the primary outcome, although, anxiety symptoms decreased over time in all conditions with only modest and non-significant differential change. None of the effects found would survive adjustments for multiple testing. There were no significant differences at follow-up for any of the outcome measures.

**Table 3.**
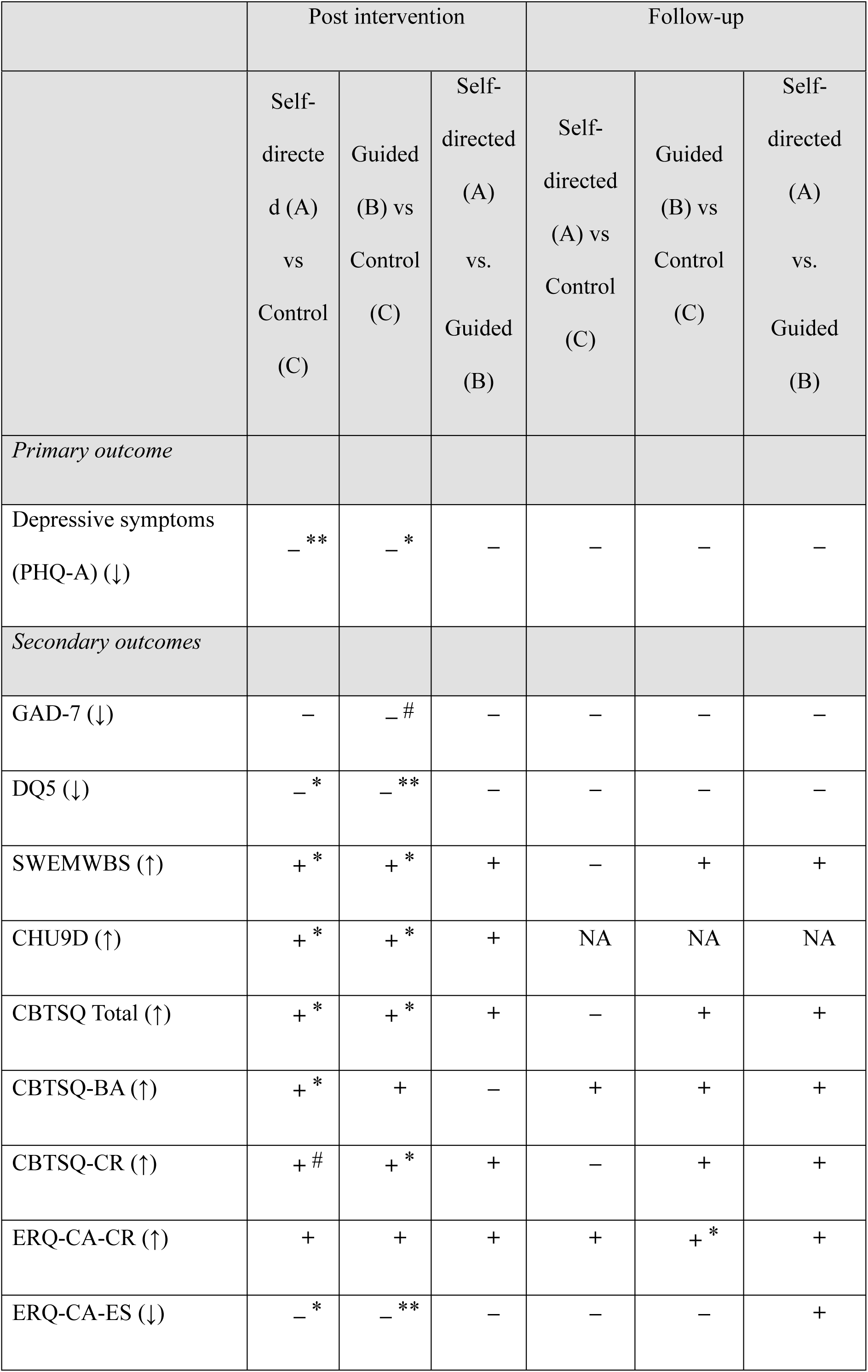
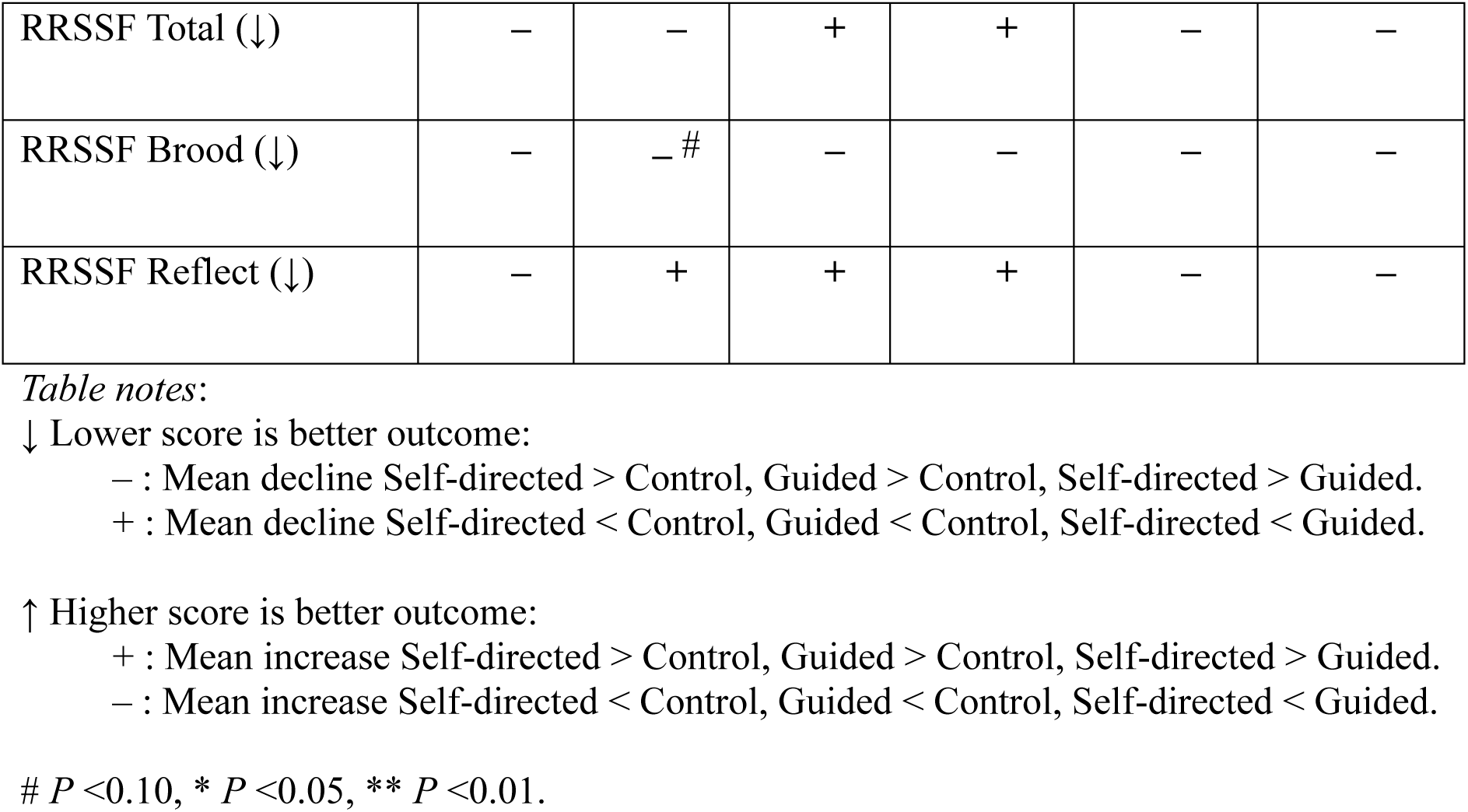
Direction of effect and statistical significance for differences in change from baseline at each occasion of measurement for each condition for primary and secondary outcomes.

There was no significant difference in the uptake of ClearlyMe^®^ between the self-directed (88.7%, *n*=173/195) and guided conditions (90.8%, *n*=167/184, OR: 1.02, 95% CI:0.96 – 1.10, *x*^2^ = 0.43, df=1, *P=*.513). Participants in the self-directed and guided conditions completed an average of 2.09 (SD: 2.82, Mdn: 1, IQR: 3) and 2.80 (SD: 2.84, Mdn: 2, IQR: 5) collections, respectively. A substantial percentage of participants in each group completed no collections (see Supplementary Material 2). Condition was a significant predictor of completing no collections (OR: 2.93, *P*=.009, 95% CI: 1.31 – 6.55) but was not associated with the total number completed (IRR: 1.03, *P*=.818, 95% CI: .80 – 1.32). Significantly more participants undertook at least one collection in the guided condition (72.5%) compared to the self-directed condition (55.8%, relative likelihood: 1.31, *P* =.001, 95% CI:1.11 – 1.54).

However, those in the guided condition (M: 3.86, SD: 2.66) did not go onto complete significantly more collections than those in the self-directed condition (M: 3.77, SD: 2.83, IRR: 1.02, *P*=.807, 95% CI: 0.85 – 1.23). Shown in the Supplementary Material 2, there was mixed evidence of a significant difference in the number of activities completed between conditions with one analysis suggesting guided (M: 13.31, SD: 9.43, Mdn: 12, IQR: 15) was superior to self-directed (M: 10.80, SD: 10.08, Mdn: 8, IQR: 15) for increasing activities completed. When examining the effects of app engagement on primary outcomes at post intervention, an expectancy effect was found as demonstrated by the emerging symptom improvements in participants who completed no activities and the small dose effect in both conditions. The guided condition reported significantly higher total digital working alliance scores than the self-directed condition (see Table 4, *t*(249)=-3.14, *P*=.002).

**Table 4.**
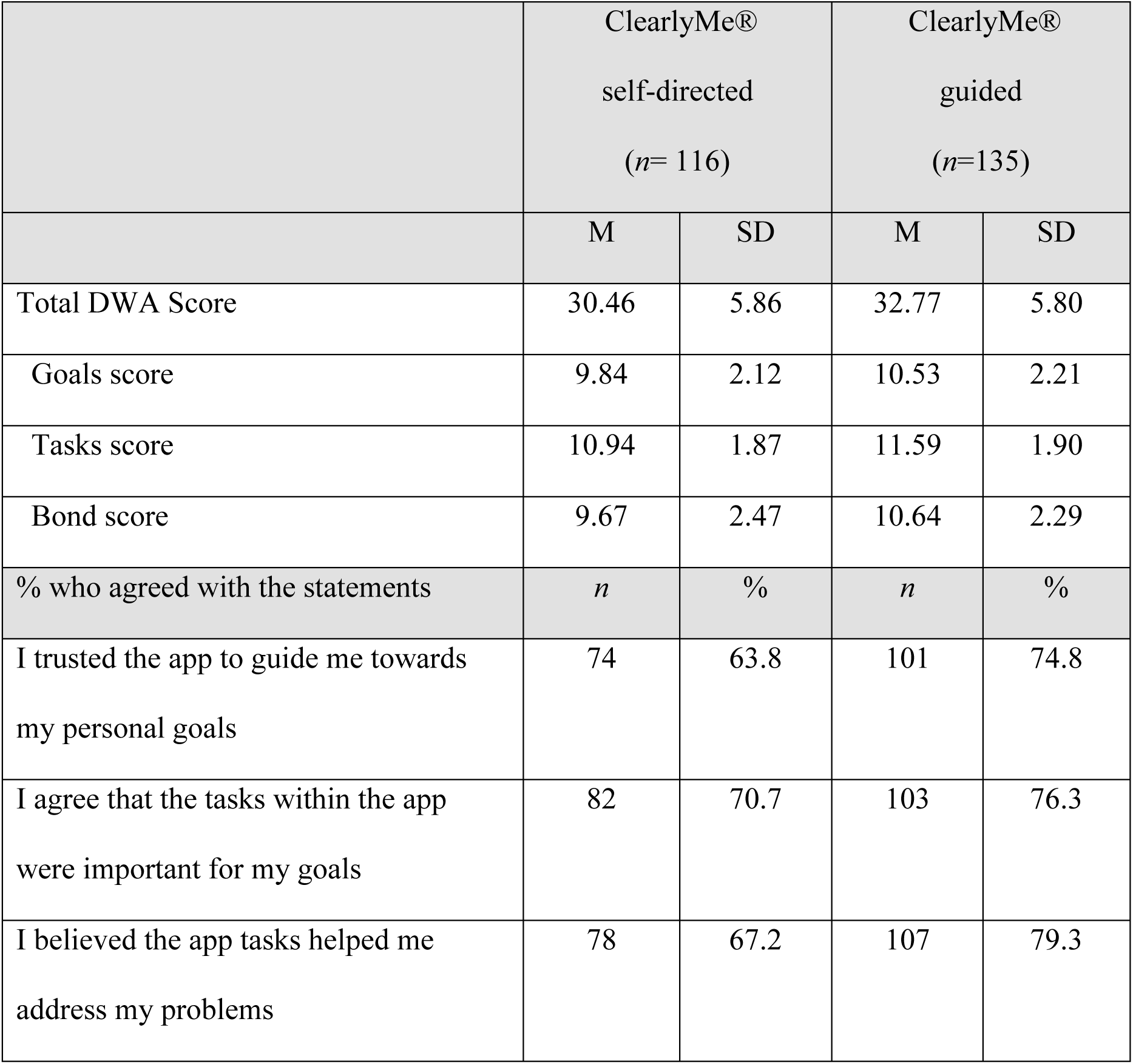
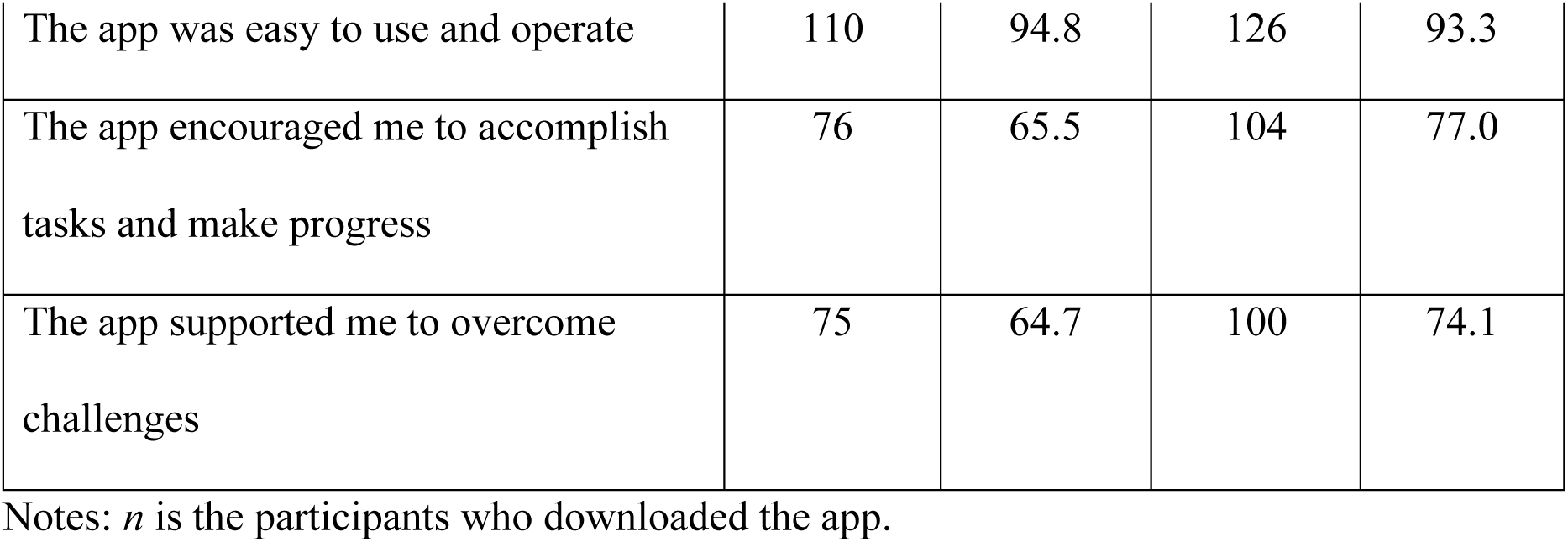
Digital working alliance (DWA) scores across the active intervention conditions post intervention.

|  | ClearlyMe®<br>self-directed<br>(*n*= 116) |  | ClearlyMe®<br>guided<br>(*n*=135) |  |
| --- | --- | --- | --- | --- |
|  | M | SD | M | SD |
| Total DWA Score | 30.46 | 5.86 | 32.77 | 5.80 |
| Goals score | 9.84 | 2.12 | 10.53 | 2.21 |
| Tasks score | 10.94 | 1.87 | 11.59 | 1.90 |
| Bond score | 9.67 | 2.47 | 10.64 | 2.29 |
| % who agreed with the statements | *n* | % | *n* | % |
| I trusted the app to guide me towards my personal goals | 74 | 63.8 | 101 | 74.8 |
| I agree that the tasks within the app were important for my goals | 82 | 70.7 | 103 | 76.3 |
| I believed the app tasks helped me address my problems | 78 | 67.2 | 107 | 79.3 |
| The app was easy to use and operate | 110 | 94.8 | 126 | 93.3 |
| The app encouraged me to accomplish tasks and make progress | 76 | 65.5 | 104 | 77.0 |
| The app supported me to overcome challenges | 75 | 64.7 | 100 | 74.1 |

Notes: *n* is the participants who downloaded the app.

High levels of satisfaction were reported across all three conditions, indicating the interventions were easy and enjoyable to use (see Table 5). Helpfulness scores were also similar (self-directed M: 3.61, SD: 0.76, guided M: 3.89, SD: 0.67, control M: 3.64, SD: 0.83). A common barrier to use was forgetfulness, with technical and user experience barriers less common (<5%). Generally, the SMS-supported condition reported fewer barriers to use than the self-directed condition. The qualitative analysis indicated that ClearlyMe^®^ supported users learning strategies to challenge unhelpful thoughts, feelings and behaviours, while providing a sense of support within an accessible and safe environment. Suggestions included greater personalisation, more inclusive content, improved navigation, reminders, and progress trackers (see Supplementary Material 2).

**Table 5.**
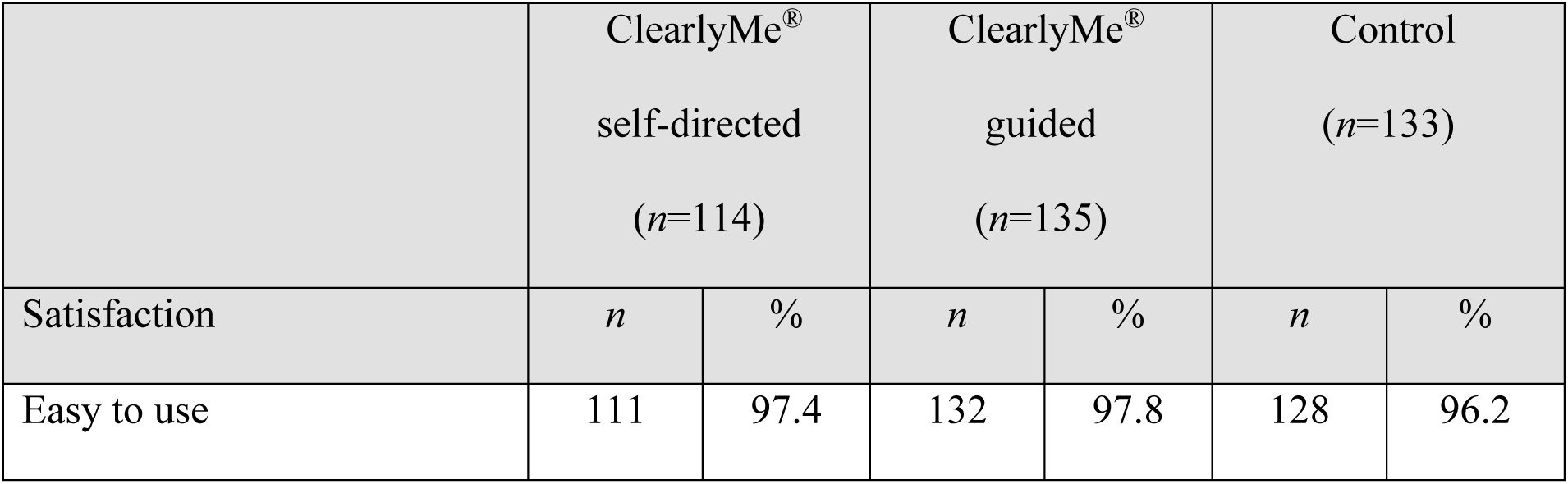
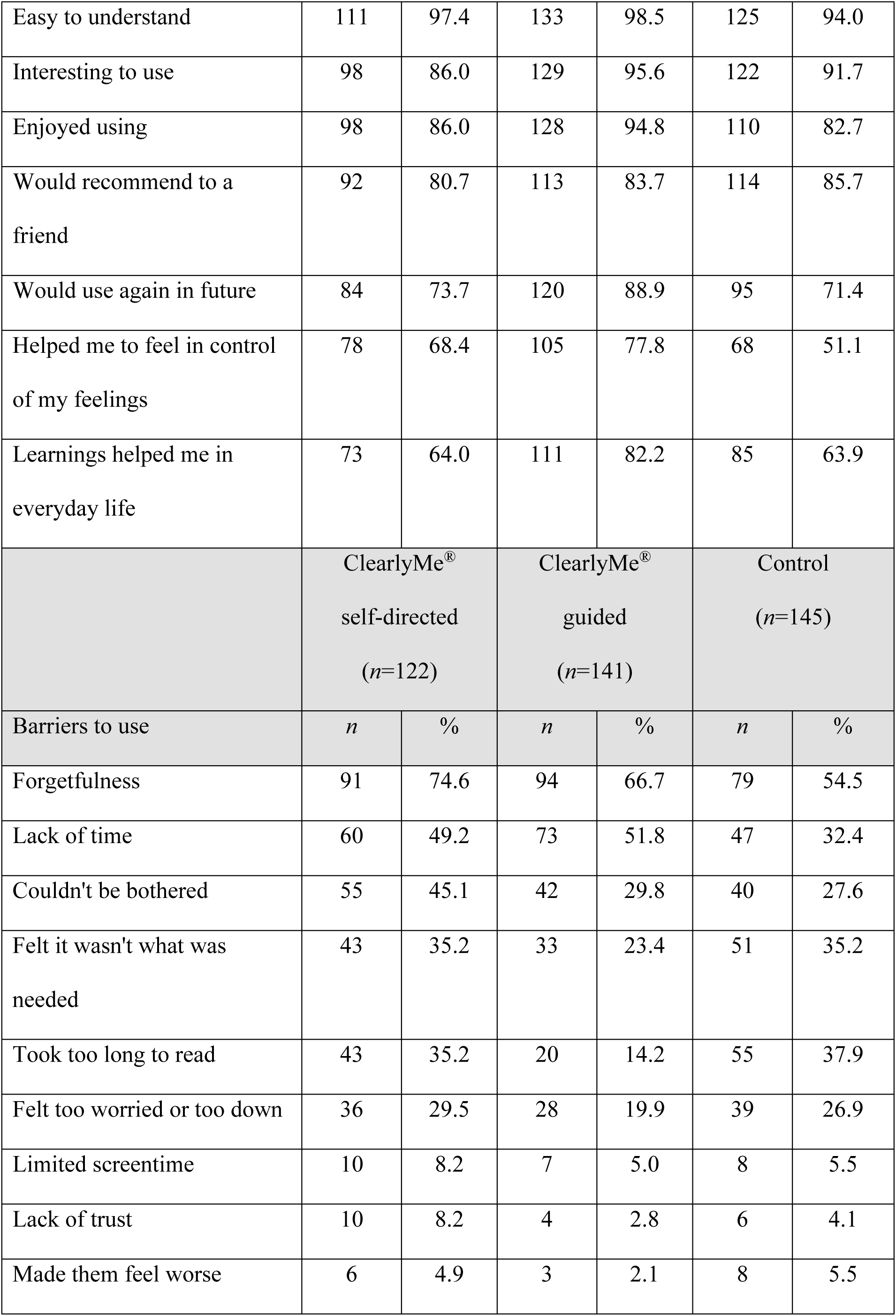
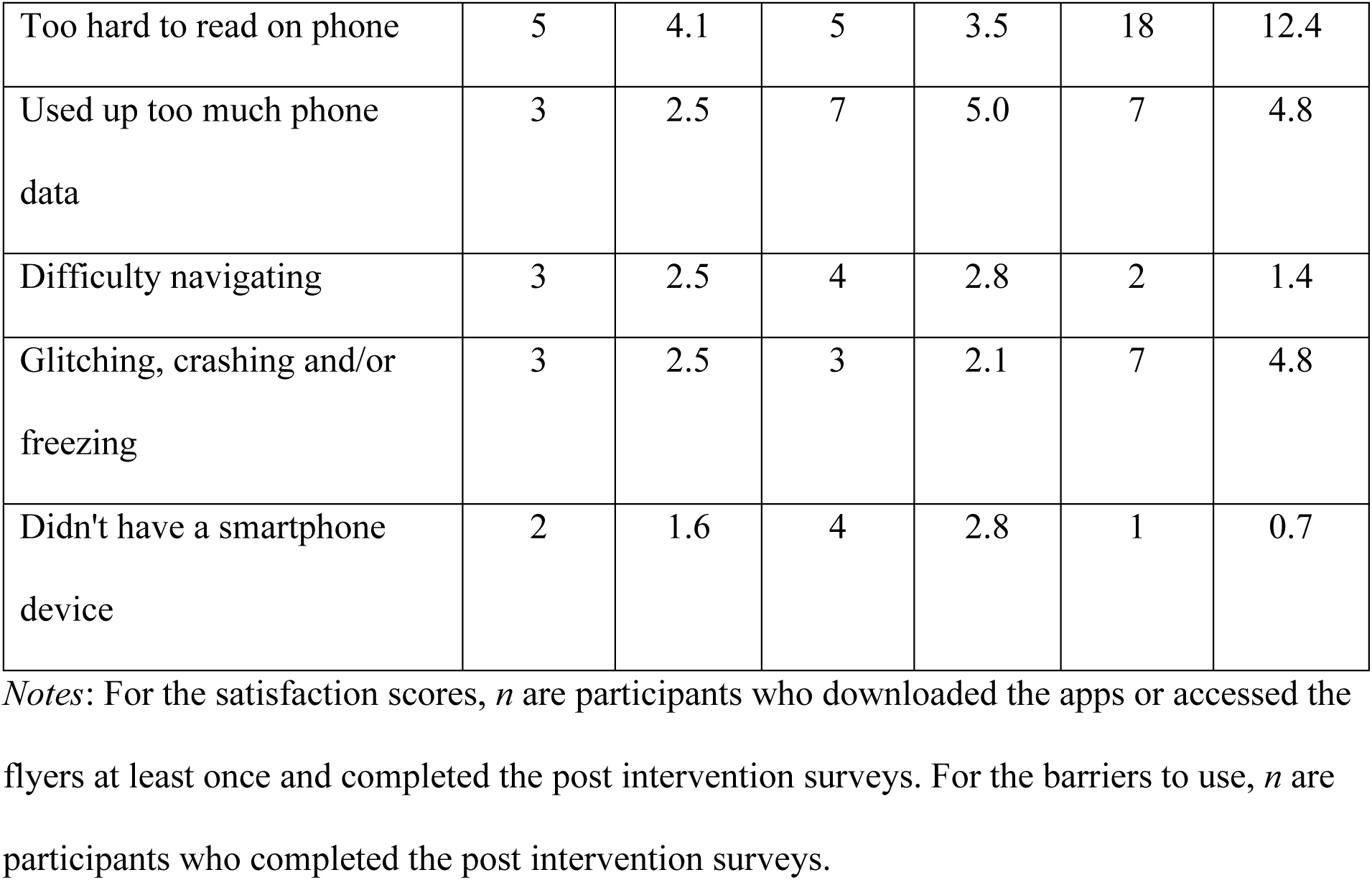
Program satisfaction and barriers to use across the three conditions.

|  | ClearlyMe<sup>®</sup><br>self-directed<br>(*n*=114) |  | ClearlyMe<sup>®</sup><br>guided<br>(*n*=135) |  | Control<br>(*n*=133) |  |
| --- | --- | --- | --- | --- | --- | --- |
| Satisfaction | *n* | % | *n* | % | *n* | % |
| Easy to use | 111 | 97.4 | 132 | 97.8 | 128 | 96.2 |
| Easy to understand | 111 | 97.4 | 133 | 98.5 | 125 | 94.0 |
| Interesting to use | 98 | 86.0 | 129 | 95.6 | 122 | 91.7 |
| Enjoyed using | 98 | 86.0 | 128 | 94.8 | 110 | 82.7 |
| Would recommend to a friend | 92 | 80.7 | 113 | 83.7 | 114 | 85.7 |
| Would use again in future | 84 | 73.7 | 120 | 88.9 | 95 | 71.4 |
| Helped me to feel in control of my feelings | 78 | 68.4 | 105 | 77.8 | 68 | 51.1 |
| Learnings helped me in everyday life | 73 | 64.0 | 111 | 82.2 | 85 | 63.9 |
|  | ClearlyMe<sup>®</sup><br>self-directed<br>(n=122) |  | ClearlyMe<sup>®</sup><br>guided<br>(n=141) |  | Control<br>(n=145) |  |
| Barriers to use | *n* | % | *n* | % | *n* | % |
| Forgetfulness | 91 | 74.6 | 94 | 66.7 | 79 | 54.5 |
| Lack of time | 60 | 49.2 | 73 | 51.8 | 47 | 32.4 |
| Couldn't be bothered | 55 | 45.1 | 42 | 29.8 | 40 | 27.6 |
| Felt it wasn't what was needed | 43 | 35.2 | 33 | 23.4 | 51 | 35.2 |
| Took too long to read | 43 | 35.2 | 20 | 14.2 | 55 | 37.9 |
| Felt too worried or too down | 36 | 29.5 | 28 | 19.9 | 39 | 26.9 |
| Limited screentime | 10 | 8.2 | 7 | 5.0 | 8 | 5.5 |
| Lack of trust | 10 | 8.2 | 4 | 2.8 | 6 | 4.1 |
| Made them feel worse | 6 | 4.9 | 3 | 2.1 | 8 | 5.5 |
| Too hard to read on phone | 5 | 4.1 | 5 | 3.5 | 18 | 12.4 |
| Used up too much phone data | 3 | 2.5 | 7 | 5.0 | 7 | 4.8 |
| Difficulty navigating | 3 | 2.5 | 4 | 2.8 | 2 | 1.4 |
| Glitching, crashing and/or freezing | 3 | 2.5 | 3 | 2.1 | 7 | 4.8 |
| Didn't have a smartphone device | 2 | 1.6 | 4 | 2.8 | 1 | 0.7 |

*Notes:* For the satisfaction scores, *n* are participants who downloaded the apps or accessed the flyers at least once and completed the post intervention surveys. For the barriers to use, *n* are participants who completed the post intervention surveys.

## Discussion

This is the first RCT that has specifically tested a CBT-based smartphone app, in self-directed and guided formats, with an attention-matched control in a sample of adolescents aged 12 to 18 years who have symptoms of depression and are not receiving any other mental health treatment. ClearlyMe^®^, whether self-directed or guided, was effective for reducing depressive symptoms in adolescents after six weeks compared to SMS-delivered psychoeducation. The overall effect sizes were small yet comparable to prior research^17^ and symptom reductions were more robust and substantially larger among those who were a probable case of depression at baseline. At six weeks, ClearlyMe^®^ also significantly reduced probable cases of MDD, psychological distress, and improved wellbeing and quality of life. No differential effects were observed between self-directed and guided formats for any secondary outcomes. This trial demonstrates that ClearlyMe^®^ can be effective for reducing depressive symptoms in adolescents who have mild to moderate depression and are not receiving any other treatment.

However, contrary to the hypotheses, no differential effects were observed between the conditions at follow-up, suggesting that sustained benefits may require additional support or intervention.

Contrary to the hypothesis, guided support did not enhance engagement with ClearlyMe^®^ as measured by collections completed. However, guided support positively impacted other aspects of engagement, including increased likelihood of completing at least one collection, higher activity completion rates, fewer reported barriers, and greater therapeutic alliance. The findings suggest that although the co-design ^34^ of ClearlyMe^®^ may have improved engagement broadly, guided support appeared to increase initial use. Understanding how to optimise adolescent engagement with self-directed mental health apps remains a priority given the low completion of ClearlyMe^®^ and poor uptake of publicly released mental health apps generally ^21,22^. Alternative engagement strategies, such as tailored motivational messaging and features targeting internal motivation, may improve self-directed uptake and use of dCBT for adolescents ^37^.

The findings can also inform dissemination strategies for ClearlyMe^®^. Smartphone apps offer a low-cost, scalable approach to mental health care, and our results support ClearlyMe^®^’s potential for public release. However, exploratory analyses highlight key considerations for future research. Males in the trial showed delayed benefits, suggesting there may be potential differences in how they engage with mental health apps ^38^. Further testing is therefore needed before ClearlyMe^®^ is specifically targeted at adolescent males. Second, not all adolescents with depressive symptoms were receptive to smartphone therapy for their wellbeing^39^ and openness to this modality may have influenced treatment effects. Enhancing adolescents’ mental health literacy and their digital therapeutic relationships through marketing materials may help improve engagement ^40^. Additionally, integrating lived experience perspectives into dissemination strategies could strengthen credibility and uptake of this approach ^41^. Finally, the absence of sustained effects at follow-up suggests that some adolescents may require additional support or intervention after using ClearlyMe^®^. Future research should explore ways to facilitate re-engagement or transition to other forms of treatment. As these conclusions are based on several secondary analyses with no adjustment for multiple testing, care should be exercised in their interpretation: replication in focused, adequately powered trials is required.

### Limitations

This trial excluded adolescents with severe suicidal ideation or recent suicidal behaviour, limiting generalisability. Given the low risk of harm observed, future studies should assess ClearlyMe^®^’s effectiveness in this group. Notably, 40% of screened adolescents were ineligible due to suicidal ideation, highlighting an unmet need for digital interventions in this population. Continued exclusion of these adolescents risk widening health disparities and limiting real-world applicability^42^. Future trials should also recruit more males, younger adolescents, and those not in school to improve generalisability. While this study include gender and sexuality-diverse youth at rates similar to prior Australian mental health trials ^36^, their representation was higher than the general adolescent population ^43^. This may reflect the increased mental health needs of this group ^43^ and the Black Dog Institute’s allyship. Finally, the lack of significant effects on anxiety symptoms contrasts with prior dCBT trials ^8^. Future studies may benefit from adopting co-primary outcomes to ensure that trials are adequately designed and powered to detect effects on anxiety, given its co-morbidity with depression.

## Conclusion

This trial demonstrated that ClearlyMe^®^, whether self-directed or guided, was effective for reducing depressive symptoms in adolescents with mild to moderate depression. Future research should evaluate its utility in underrepresented groups, including adolescents with suicidality, males, and those not in schooling. Implementation efforts should consider adolescents’ openness to digital mental health interventions to maximise impact.

### Contributors

BOD, AWS, HC, MRA, MSK conceived the project and were awarded funding. SHL, MRA, MSK and BOD designed the intervention. BOD, AWS, HC, PJB, AJM, SHL, MRA, MSK, PJB and AJM designed the trial and study protocol. SHL, MSK, MRA, AR coordinated trial recruitment and trial operations including the provision of guided and technical support, overseen by BOD and AWS. SHL and AWS provided clinical supervision to the trial. AJM was the trial statistician and conducted all analyses. MRA, MSK, KN, ZD, BG conducted the qualitative analyses. BOD, MRA and MSK prepared the manuscript, and all authors reviewed.

### Data sharing

De-identified participant data and a data dictionary defining the relevant variables can be made available to others upon request by contacting the Chief Investigator and with all new personnel modifications approved by the University of New South Wales Human Research Ethics Committee, prior to data sharing. Trial related documents including the protocol and informed consent forms are publicly accessible on the Australia and New Zealand Trial Registry.

## Data Availability

All data produced in the present study are available upon reasonable request to the authors.

## Acknowledgements.

The MobiliseMe study was supported by philanthropic funding from the Goodman Foundation and the National Health and Medical Research Council (NHMRC) Investigator Grants (BOD grant number MRF1197249, AWS grant number: GNT2008839, HC grant number GNT115614). The authors would like to acknowledge the involvement of the adolescents in the trial. The authors would also like to acknowledge the Trial Data Safety and Monitoring Board members including Professor Jill M. Newby, Associate Professor Alexis E. Whitton, Associate Professor Michelle Tye, and Professor Andrew J. Mackinnon. The authors also acknowledge Dr Ashleigh Ticknell for providing administrative support in the guided use condition, as well as the clinical psychologists, Dr Samantha Tang, Dr Joanne Beames, Emily Upton and Britt Corkish who assisted with the trial safety protocols.

