## Supplementary Material 1 for "The MobiliseMe study: A randomised controlled efficacy trial of a cognitive behavioural therapy smartphone application (ClearlyMe®) for reducing depressive symptoms in adolescents"

Black Dog Institute

### Supplementary Material 1

### Overview of ClearlyMe®

#### ClearlyMe®

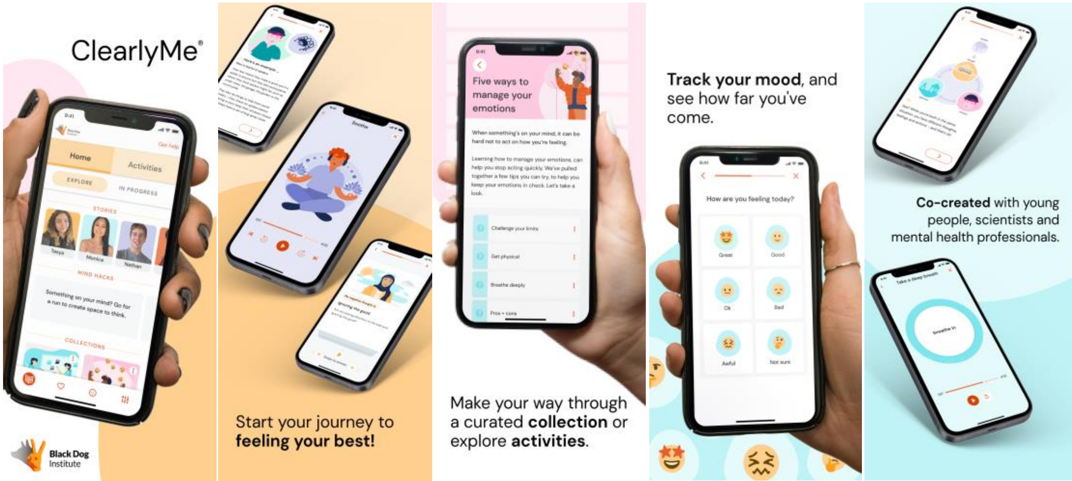


The first version of ClearlyMe® was used for the MobiliseMe trial in 2022 and details of this version are outlined below. The app was released publicly in June 2024 and included changes to the onboarding. At the time of this publication the content within ClearlyMe® has remained the same.

**ClearlyMe: Version one (used in the MobiliseMe RCT)**

ClearlyMe is a free, self-directed CBT-based smartphone application that provides therapeutic content and symptom management strategies to adolescents experiencing mild to moderate depressive and anxiety symptoms. The application was specifically designed for adolescents (aged 12-17 years) by a multidisciplinary team of psychologists, researchers, user experience designers and software developers at the Black Dog Institute. ClearlyMe is delivered as an autonomous, non-sequential CBT program consisting of 37 brief lessons (i.e., therapeutic content alongside activities). Lessons include the following evidence-based treatment components: psychoeducation, cognitive restructuring, emotion awareness and acceptance, goal setting, problem solving, activity scheduling, behavioural activation, exposure, relaxation, mindfulness, and values labelling.

The lessons also encourage participants to practise the psychological skills in-between lessons and return to the app to reflect or access further content as needed. The lessons vary in length, taking between 5 to 10 minutes each to complete. To help navigate the program content, users are encouraged to complete the lessons via nine curated 'collections'. A collection is defined as a structured selection of related lessons grouped together with a brief title and introduction. Each collection varies in length, taking approximately 20 minutes to complete. Users can also complete the individual lessons from a 'Activities' list whereby they self-select lessons considered to be relevant to their individual needs. In this list, lessons are categorised into three groups depending on their target: 'emotions', 'thoughts', 'behaviours'. The app also includes a Moodcheck (i.e., brief mood monitoring), Mind Hacks (i.e., quick strategies that help in the moment), and Stories (i.e., short videos of young peoples' experience managing mental health symptoms and positive help-seeking experience) to provide users with additional pathways to accessing therapeutic content. For these features, specific lessons are recommended after the feature is accessed. ClearlyMe also includes in-app reminders, 'saving' and 'favourite' functions to support users to return to the app to reengage in content.  In-app reminders include a 'revisit the app' reminder, which notifies all participants to use the app if they haven't done so in the past 7 days. This reminder is locked and cannot be turned off in the app. Users can also set an additional reminder to use the Moodcheck feature once a day for as many days as they wish. The app also includes a 'Get Help' section that contains information of when and where to access additional mental health support services (e.g., Kid's Helpline).

**Table 1. Summary of ClearlyMe collections**

| Collection | Supports adolescents to… |
|---|---|
| 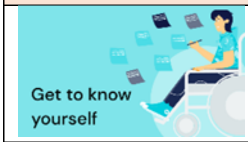  | • learn what emotions, thoughts, and behaviours are and how they work<br>• understand their own emotional triggers and develop greater control over their actions. |
| 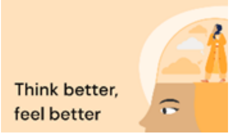  | • identify and appraise their thoughts,<br>• recognise common thinking errors,<br>• understand the link between thoughts, feelings and behaviours,<br>• create healthier thinking patterns, learn ways to challenge their unhelpful thinking habits. |
| 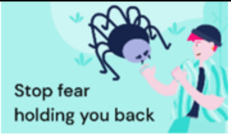  | • identify symptoms of anxiety,<br>• use a breathing exercise when they need to relax.<br>• plan steps to overcome a particular fear,<br>• recognise realistic vs unhelpful thoughts,<br>• recognise early signs when they're not coping,<br>• create a plan to deal with difficult times in the future. |
| 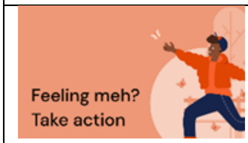  | • identify symptoms of depression,<br>• set goals, identify pleasurable activities, and develop positive habits,<br>• conduct a thought challenging experiment to test whether thoughts result in true predictions,<br>• recognise early signs when they're not coping,<br>• create a plan to deal with difficult times in the future. |
| 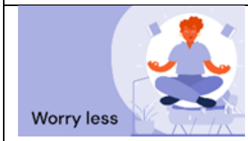  | • notice if overthinking is becoming a problem and explore whether their thinking patterns are helpful,<br>• learn how to problem-solve, weigh up the pros and cons of solutions, and practice in their daily lives,<br>• schedule worry time,<br>• learn about and practice mindfulness,<br>• set goals, identify pleasurable activities, and develop positive habits. |
| 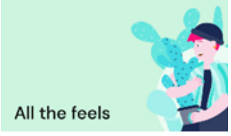  | • learn what emotions are and how they work,<br>• recognise, understand, and accept uncomfortable emotions,<br>• practice a grounding exercise to manage their emotions or thoughts,<br>• practice a mindfulness exercise. |

| | |
|---|---|
| 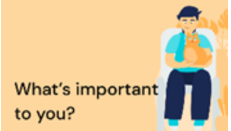  | • find out what they value to help focus on and be guided by what's important to them, and<br>• learn and practice goal-setting skills to feel more motivated and productive. |
| 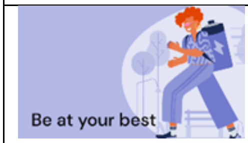  | • practice a relaxation exercise to unwind and sleep well,<br>• learn tips to stop procrastinating,<br>• practice physical exercise to improve their mood, and<br>• learn strategies to improve their self-confidence and build a plan to try. |
| 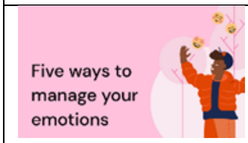  | • practice physical exercise to improve their mood,<br>• manage their emotions with a breathing exercise,<br>• problem solve with a pros and cons list and practice in their daily life,<br>• use opposite action to handle difficult emotions. |

**ClearlyMe: Version two (released publicly)**

Version 2 of ClearlyMe® is available to download on the app stores

- Google Play: https://play.google.com/store/apps/details?id=au.org.blackdoginstitute.tt&hl=en&gl=US&pli=1
- Apple App store: https://apps.apple.com/au/app/clearlyme/id1550213032

More info on The Black Dog Institute website: https://www.blackdoginstitute.org.au/research-projects/youth-cbt/

Black Dog Institute
