## Supplementary Material 2 for "The MobiliseMe study: A randomised controlled efficacy trial of a cognitive behavioural therapy smartphone application (ClearlyMe®) for reducing depressive symptoms in adolescents"

### Results

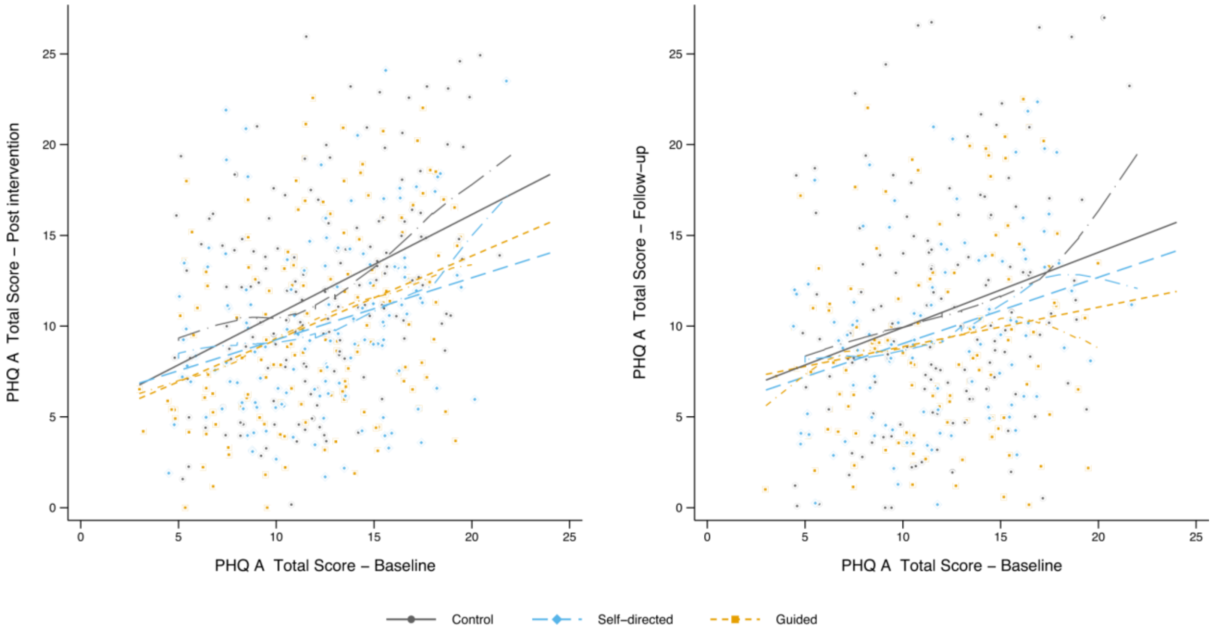

**Supplementary Figure S1. Scatterplots, linear and LOWESS fit lines for post intervention (primary endpoint) and follow up (secondary endpoint) assessments of PHQ-A against baseline status.**

Figure Note: Dashed lines: linear fit; Dash-dotted lines: LOWESS curves

*Post-hoc sensitivity analysis for changes in PHQ-A adjusting for attrition*

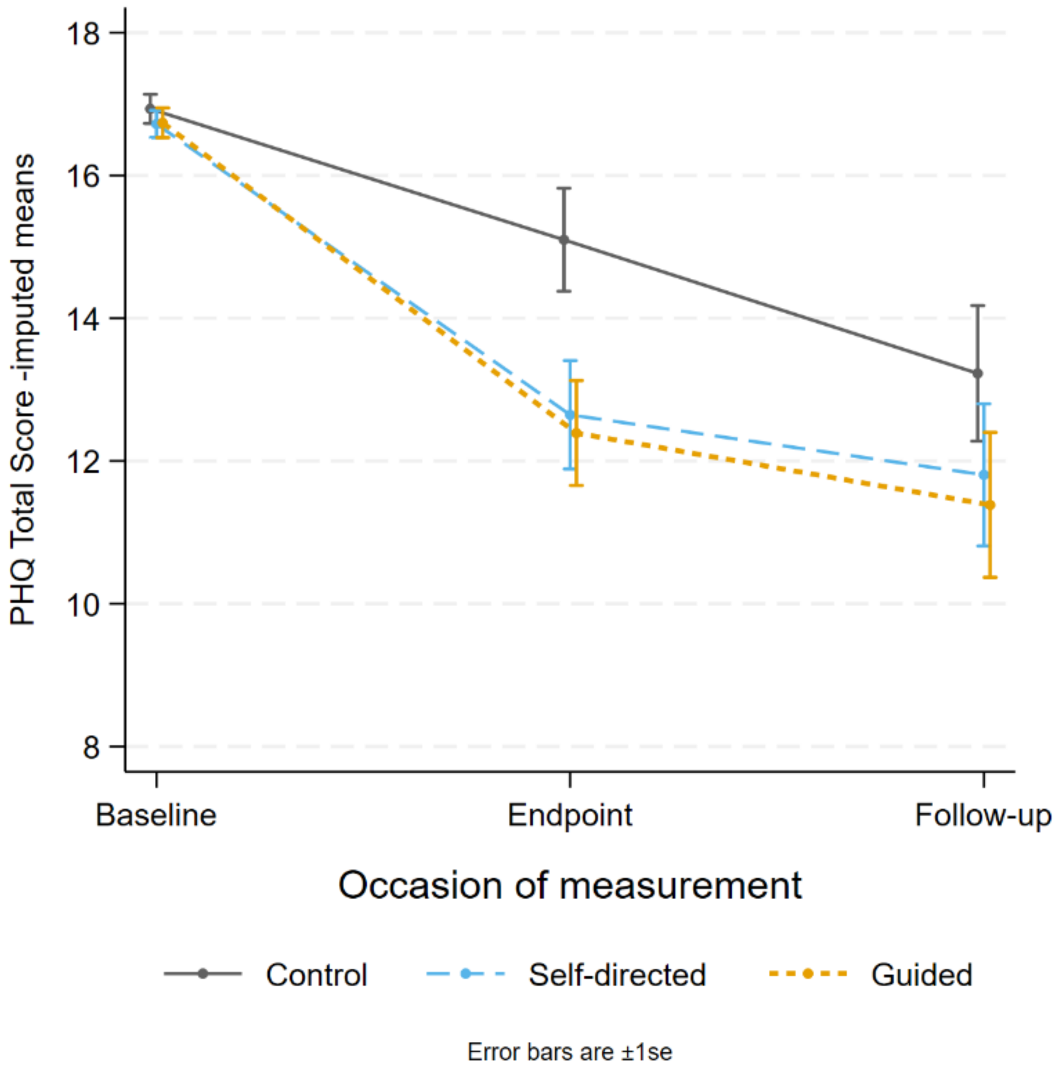

**Supplementary Figure S2. Post hoc sensitivity analysis adjusting for attrition by treating missing cases as control cases, showing reduced but persisting intervention effects on PHQ-A at primary endpoint.**

*Exploratory analysis for gender effects*

The exploratory analysis for gender effects (males and females) on depressive symptoms yielded similar patterns as the whole sample (see Supplementary Material for results).

However, there was low power for detecting gender effects given so few males in the sample (*n*=99). Among males (*n*=99), change in depressive symptoms post intervention did not significantly differ between conditions. For males, the control condition remained stable but the guided and self-directed conditions declined over time. Cross-sectionally, this difference was nearly significant at follow-up ($P$=.053) with a moderate effect size of $d$=0.58 (95%CI: -0.001 – 1.16) for guided vs control and significant ($P$=.026) for self-directed vs control with a large effect size ($d$=0.70, 95%CI: 0.09 – 1.31).

*Exploratory effects for openness to smartphone applications for mental health and wellbeing*

In participants with moderate to high openness to smartphone therapy at baseline (Figure S3), change in depressive symptoms at post intervention was significant in the self-directed and guided conditions ($P$=.009, $d$=0.34, 95%CI: 0.06 – 0.63 and $P$=.013. $d$=0.39, 95%CI: 0.11 – 0.66 respectively) but remained significant at follow-up in the guided condition only ($P$= .015, $d$=0.42, 95%CI: 0.12 – 0.71). In those with low openness at baseline (Figure S4), there were no significant changes in depressive symptoms at any time point in the self-directed or guided conditions when compared to the control (all $P$ >.05), although, symptoms in the self-directed condition declined at follow-up in comparison to means in the guided condition, which converged to baseline ($P$=.051).

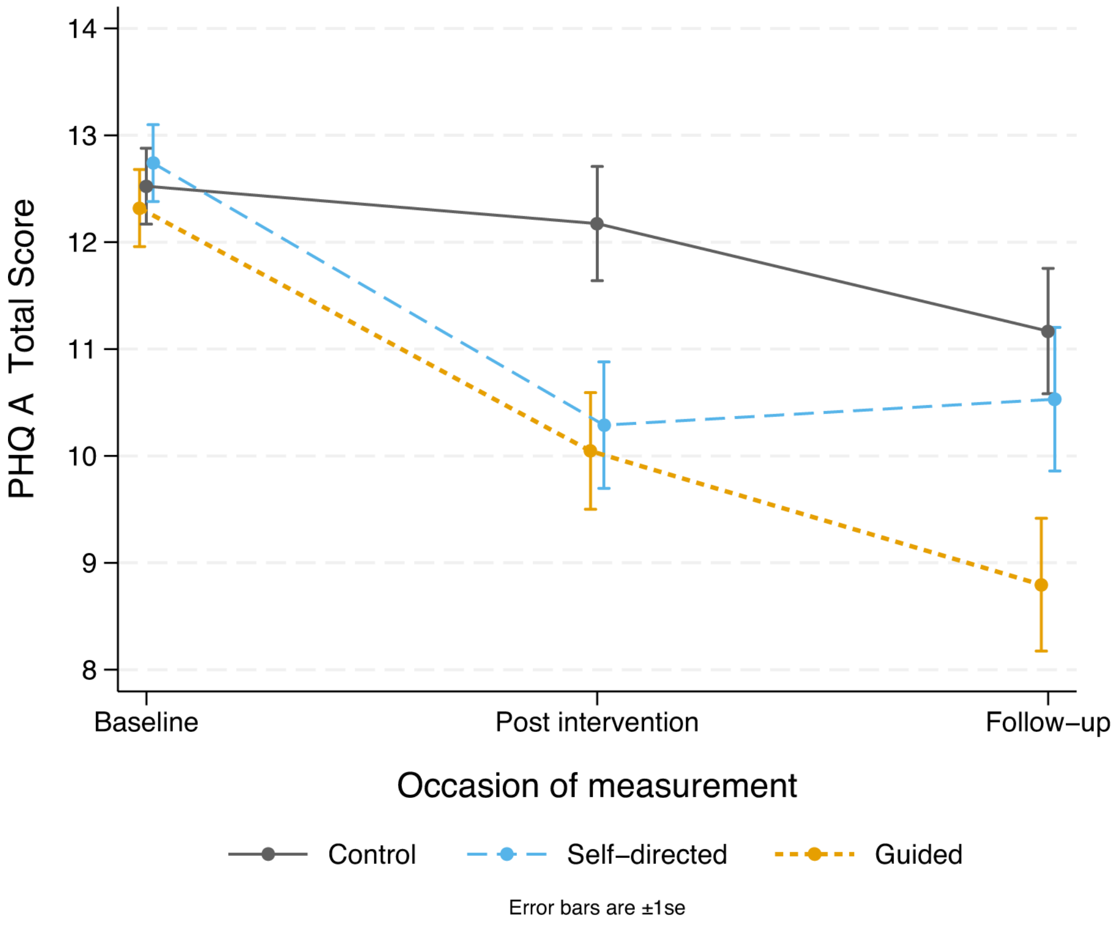

**Figure S3. Reductions in depressive symptoms (PHQ-A) among participants with moderate to high levels of openness to smartphone therapy at baseline for the conditions at each occasion of measurement.**

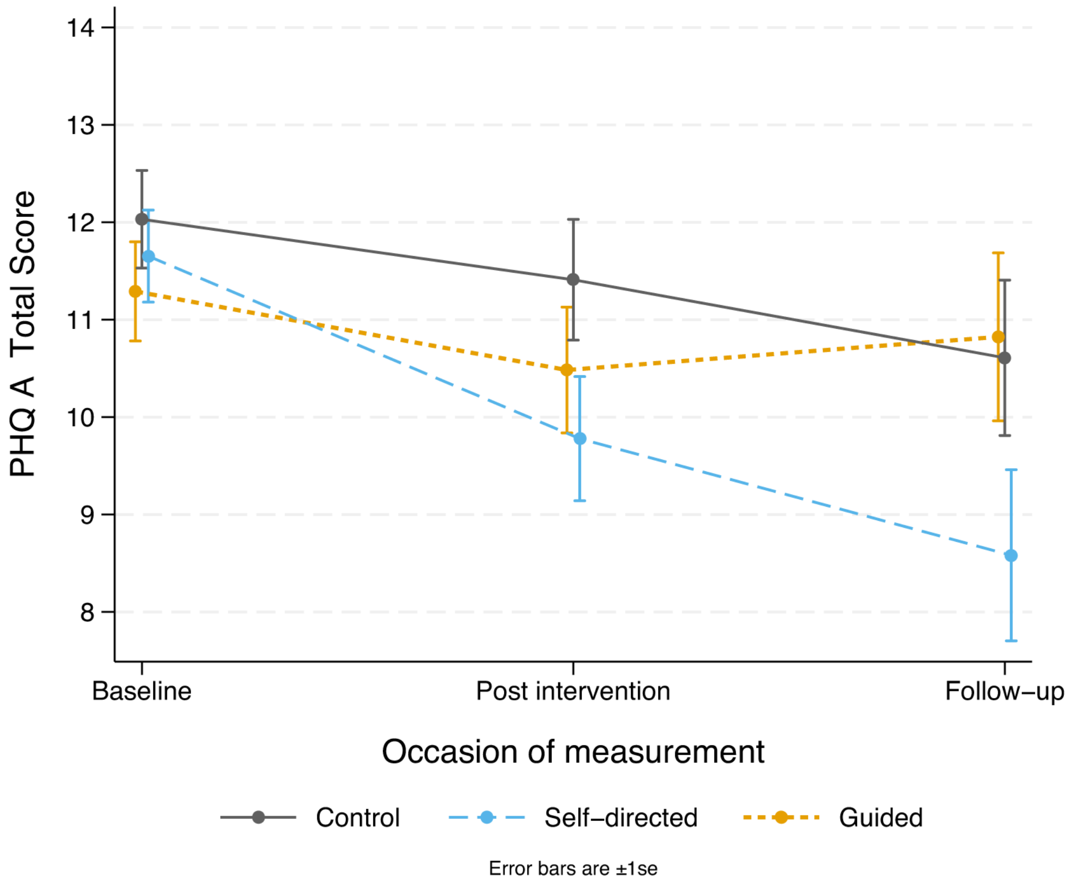

**Figure S4. Reductions in depressive symptoms (PHQ-A) among participants with low levels of openness to smartphone therapy at baseline for the conditions at each occasion of measurement.**

*Differences in collections completed across intervention conditions*

The distributions of the total number of collections completed in each active condition are shown in Figure S5.

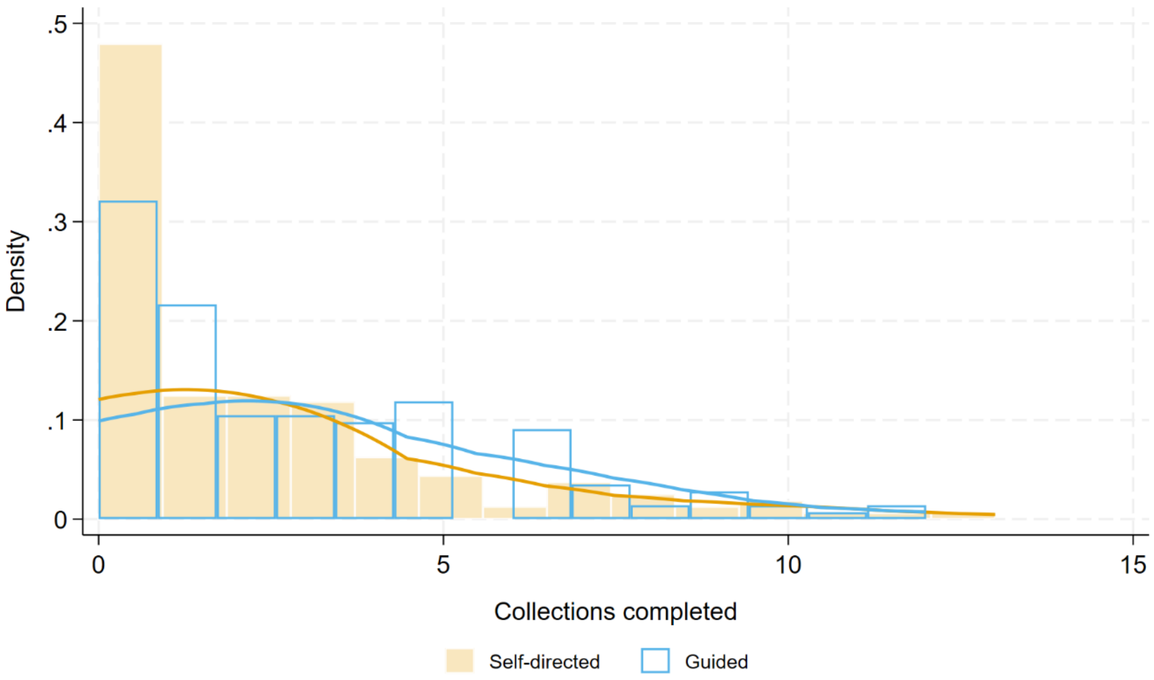

**Supplementary Figure S5. Histogram of total collections completed in ClearlyMe® self-directed and ClearlyMe® guided conditions.**

Figure note: Lines are kernel densities – 'smoothed' histograms.

*Differences in activities completed across intervention conditions*

The distributions of the total number of unique activities completed in each active condition are shown in Figure S6. Participants in the self-directed condition and the guided condition completed an average of 10.80 (SD 10.08, Mdn: 8, IQR: 15) and 13.31 (SD 9.43, Mdn: 12, IQR: 15) unique activities, respectively. A series of models of increasing complexity explored the relationship between conditions (ClearlyMe® self-directed vs ClearlyMe® guided) on the number of activities completed. A zero-inflated negative binomial model with a constant inflation component (i.e., not a function of condition) fitted the data better than Poisson and negative binomial models without zero inflation. Allowing inflation to differ between conditions did not significantly improve model fit. The significance of the effect of condition was extremely equivocal and dependent on the method of estimation. Using standard estimates, there was no significant difference between conditions (*P*=.071); however, bootstrap estimation led to a significant IRR of 1.18 (95% CI:1.00 – 1.39) and the difference in the median was significant ($x^2$ = 4.98, df=1, *P*=.026). The latter may be a more robust test of whether one group completed more activities than the other. A total of 14.5% of participants in the self-directed condition completed no activities compared to 9.0% in the guided condition and this difference was not statistically significant.

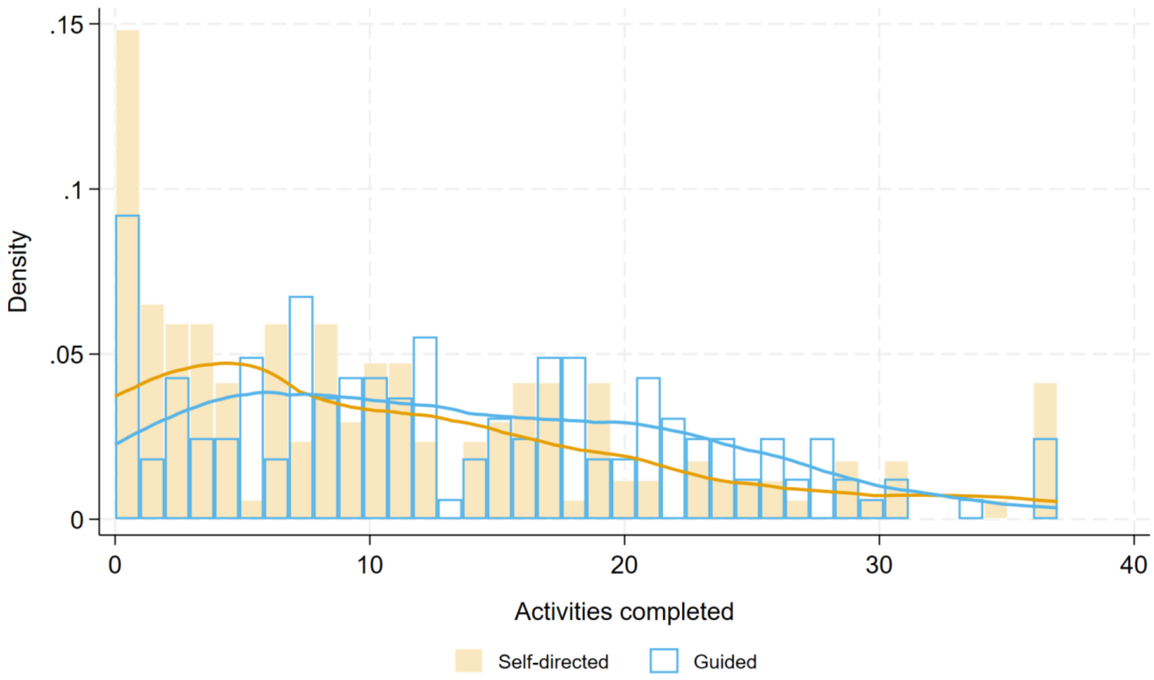

**Supplementary Figure S6. Histogram of total unique activities undertaken in the ClearlyMe® self-directed and ClearlyMe® guided chat) conditions**

*Figure note:* Lines are kernel densities – ‘smoothed’ histograms.

*Relationship between Total Unique Activities and Total Collections completed*

As can be seen in Figure S7, there was a strong relationship between Total Unique Activities and Total Collections completed within ClearlyMe®. This did not differ between conditions. The overall correlation between variables was 0.887.

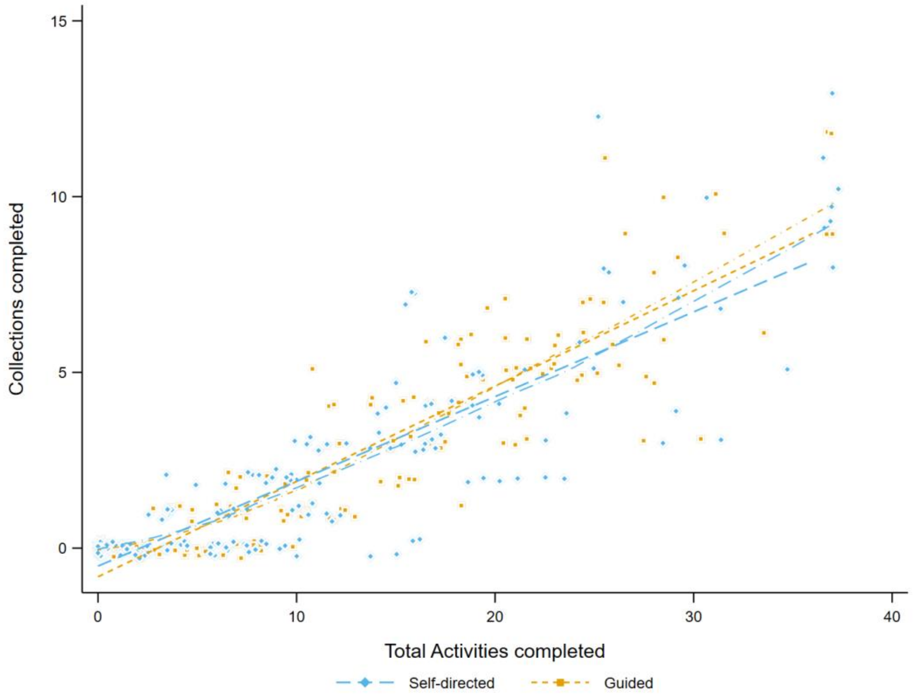

**Supplementary Figure S7. Scattergram of Total Unique Activities and Total Collections completed within ClearlyMe® self-directed and ClearlyMe® guided conditions.**

*Figure note:* Solid lines are linear relationships, dashed lines are lowess curves. Points are jittered around their actual values for clarity.

*Exploratory analysis on the potential effects of app engagement on the primary outcome.*

Mixed model repeated measures with engagement moderation analyses were conducted. Total unique activities undertaken were classified into low (fewer than 12) or high (12 or more);

this division corresponding to the median for participants who undertook at least one activity. Separate models for the PHQ were rerun including all participants in the control group and each subgroup. An additional model included only participants in the active groups who completed no activities. These participants effectively received the same 'dose' of each intervention as the control group. Outcomes of these analyses are shown in Figures S8, 9,10. Changes in depressive symptoms in the 'no activities' subgroups were somewhat greater than the control although the small samples (*n*=25 self-directed, *n*=15 guided) meant that these changes were not significant. The 'nil to low' activity subgroups were larger in sample size (*n*=84 self-directed and *n*=66 guided) and showed substantial but non-significant differential declines in depressive symptoms due, in part, to small baseline differences. Change in depressive symptoms at follow-up in the self-directed condition approached significance (*P*=.079). Cross-sectionally, post-intervention effect sizes were significant or nearly so (control vs self-directed: d=0.28, *P*=.064; control vs guided use: d=0.36, *P*=.027). The high activity subgroups (*n*=86 self-directed, *n*=103 guided use) showed a similar pattern to the low activity group, although baseline to post-intervention change was slightly larger and significant for the self-directed condition (*P*=.010) and slightly smaller but nearly significant (*P*=.079) in the guided group. Cross-sectional post-intervention effect sizes were significant in both conditions (control vs self-directed use: d=0.38, *P*=.009; control vs guided: d=0.30, *P*=.020). The overall pattern from the three analyses hints at expectancy/placebo effects (no activity subgroup and a small 'dose' effect in both conditions). There is no evidence of different effects in the active conditions. It must be remembered that the subgroups formed for these analyses are no longer random: it may be that participants who experienced greater benefit from the intervention choose to complete more activities, so causal status is uncertain.

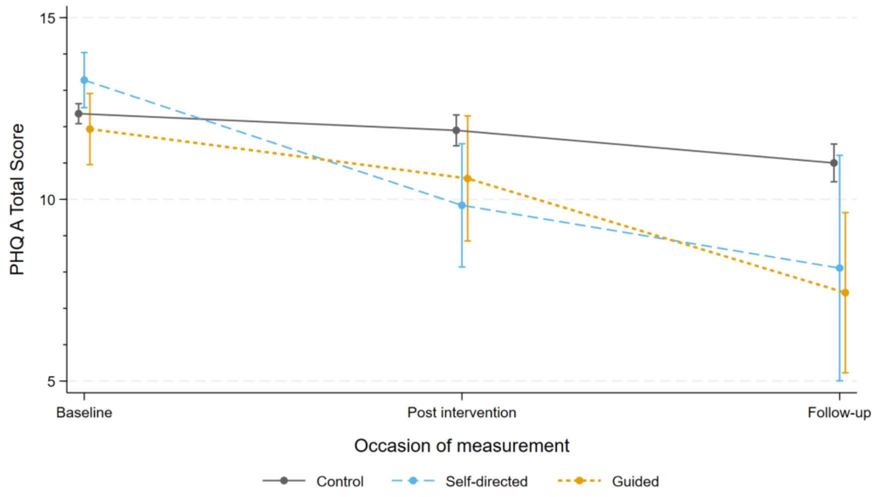

**Supplementary Figure S8. Estimated mean PHQ-A scores by condition and occasion of measurement for those completing no activities**

*Figure note:* Error bars represent ±1 se.

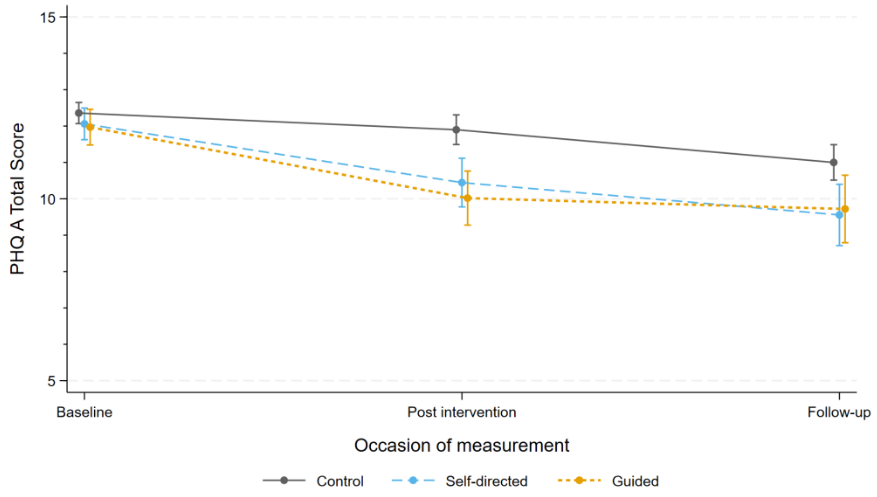

**Supplementary Figure S9. Estimated mean PHQ-A scores by condition and occasion of measurement for those completing 11 or fewer activities**

*Figure note:* Error bars represent ±1 se.

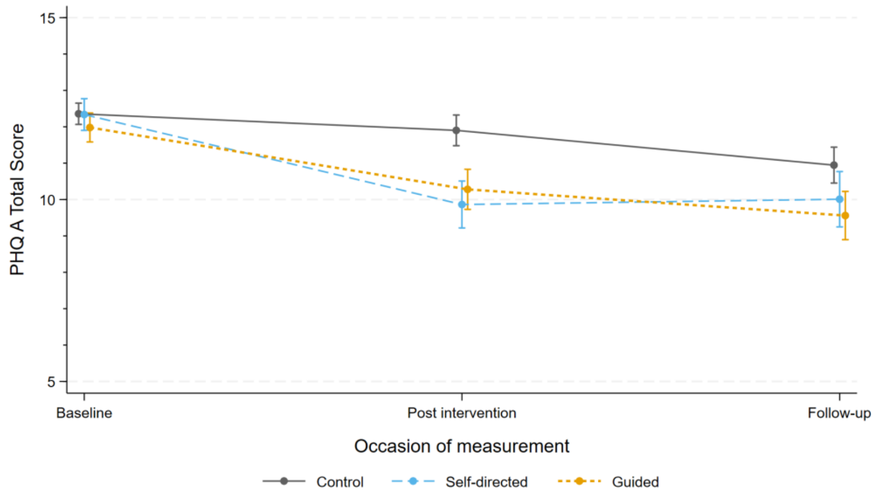

**Supplementary Figure S10. Estimated mean PHQ-A scores by condition and occasion of measurement for those completing 12 or more activities**

*Figure note:* Error bars represent ±1 se.

*Qualitative feedback on the intervention conditions.*

**Supplementary Table S1. Thematic analysis of participants' feedback on the ClearlyMe® app**.

| Themes | Description | Exemplar quote (Gender, age) |
|---|---|---|
| Strategies | Responses that described how participants found the app useful to learn strategies to identify, understand, track, and manage unhelpful thoughts, feelings, and behaviours. | "Helped me think about my thoughts and feelings and how to understand them." (F, 12-14 years)<br>"Helped me create effective management strategies when I felt down." (F, 12-14 years) |
| Outcomes | Responses that described how participants had found the app changed their thoughts, feelings, and behaviours, and found it helped with motivation, overcoming fear, relationships and self-esteem. | "It was almost meditative in a way. The voices were calming, and I used it on my commute when my thoughts were racing, and it grounded me." (M, 15-17 years)<br>"Honestly, the times I used it, got me motivated that I was actually making an improvement." (M, 12-14 years)<br>"Something for me to do when I was feeling really anxious." (GF, 15-17 years), "Just having a little time every day to focus on myself has helped me detach." (F, 15-17 years). |

| | | |
|---|---|---|
| | Responses that described ways the app could be changed to improve outcomes, as it was a source of distraction or distress. | "I feel that some of the modules made me feel worse because I had to identify flaws about myself." (GQ, 15-17 years)<br><br>"Not send messages to make me feel like I have to complete them as it stresses me out." (F, 15-17 years) |
| Accessibility | Responses expressing the strengths of the app, including providing a safe opportunity, space, time, and/or engaging platform to reflect and experiment with learned strategies. | "It provided me with a platform to focus on my mental health." (F, 15-17 years )<br><br>"It gave me a place to work through the type of activities that counsellors have suggested in the past but are hard to initiate." (NB, 15-17 years ) |
| | Responses expressing how the app could be improved, as | "More recognition of how young people feel and how simple thinking strategies won't fix everything." (M, 15-17 years) |

| | | |
|---|---|---|
| | participants found it was not engaging or inclusive. | "I'm not sure but it just doesn't keep my interest it felt like homework." (F, 12-14 years) |
| Support | Responses showing that participants' use of the app made them feel supported, including the "Stories" feature. | "I made me realise that I wasn't alone with my struggles and that there are things/people out there who truly want to help me." (F, 15-17 years)<br><br>"…being able to see other people's stories made me feel connected, relatable and supported" (M, 12-14 years)<br><br>"the peoples stories reminded me I wasn't alone." (F, 15-17 years )<br><br>"I really enjoyed the stories so I think it would be good if there were more of them." (NB, 15-17 years) |
| | Responses showing use of the app made them feel abandoned | "Advice instead of just making me acknowledge my feelings and then stopping go and leaving me in a bad place with no way to understand how to control what the app made me address." (F, 15-17 years) |
| App experience | Responses which suggested there were some technical issues and suggested | "General bug fixes." (F, 15-17 years ), "Notifications because I forgot sometimes." (F, 15-17 years ) |

| | | |
|---|---|---|
| | improvements to the app, including adding notifications, a progress tracker, FAQ's/forum, clearer layout, visual design, plus additional features (e.g. gamification, journal, calendar) | "It would be good if this app also had games to play for each mood." (F, 12-14 years)<br><br>"A progress bar so I can see how far through a set I am." (F, 15-17 years)<br><br>"the colours of the app…need to be changed." (NB, 15-17 years) |
| Content variety | Responses expressing a desire for more activities and/or collections. | "Adding a few more activities." (M, 15-17 years), "I would add more collections for people with different types of problems, as I feel that there aren't enough collections." (F, 15-17 years) |
| | Responses expressing a desire for more activities and/or collections. | "Make more short activities. And less collections, they were awful." (NB, 15-17 years) |
| Content personalisation | Responses that express desire for content and the user experience to be improved through personalisation, relevance and complexity. | "More specialised collections, some shorter/long or easier/harder depending on the head space sometimes I need a quick easy one to help and am too impatient to commit to a full one." (F, 15-17 years)<br><br>"The activities didn't address the problems that I am having." (NB, 15-17 years) |

| | | |
|---|---|---|
| | | "Easier activities that are less draining to think about." (F, 12-14 years)<br><br>"It was quite basic, and while it did help to some degree, I found that the activities lacked depth and could be developed further to allow for deeper thinking." (M, 15-17 years)<br><br>"Like a repeated activity instead of teaching and let go of student. (F, 15-17 years)" |

*Table Note.* F: Female. M: Male. NB: Non-binary

**Supplementary Table S2. Thematic analysis of participants' responses regarding the psychoeducation flyers**

| Theme | Description | Exemplar quote (Gender, age) |
|---|---|---|
| Content | Responses showing content was helpful or sufficient | "Helped me learn mental health literacy." (NB, 15-17 years)<br><br>"Relevant to my life." (F, 15-17 years) |
| | Responses expressing further depth or support required | "Exercises… that can be done, like what you would get with a professional, to complete, especially for those who can't access treatment, or are on a (usually very) long waiting list." (NB, 15-17 years) |

| | | |
|---|---|---|
| | | "Personal stories." (F, 15-17 years)<br><br>"Reassurance that it will be okay and everyone's different." (M, 15-17 years) |
| Outcomes | Responses expressing the psychoeducation flyers supported participants to change feelings and behaviours, including support, motivation, self-management and relationships | "Gave me hope and encouragement that I can get through this and showed me how to cope and improve" (M, 15-17 years)<br><br>"by motivating me to be my best self and gave me tips and tricks on how I can handle anxiety better." (F, 15-17 years)<br>"It helped me manage my mental health." (F, 15-17 years)<br><br>"Helped me understand mental health in myself and those around me and also recognising unhealthy friendships." (F, 15-17 years) |
| Engagement | Participants suggested the flyers could be more engaging and relevant, specifically in the formatting | "Maybe videos instead because they're more engaging." (F, 15-17 years)<br><br>"Better formatting and aesthetics... I am more likely to read and understand information that is delivered in an easy to read, professional way." (M, 15-17 years) |

| | | |
|---|---|---|
| | | "More relatable for teenagers." (F, 15-17 years) |
| Accessibility | Responses relating to the accessibility of the flyers | "They were really easy to always come back to when I needed them." (F, 15-17 years) |
| | Responses suggesting that accessibility, interoperability and availability could be improved | "Making them more fun or accessible in other locations such as Instagram." (F, 15-17 years)<br>"If they were more easily accessible and can be more easily shared." (F, 15-17 years) |

*Table Note.* F: Female. M: Male. NB: Non-binary. Age ranges provided to protect participants' privacy.
